# APOE ε4 Modifies the Association of Left Atrial Remodeling With Brain Aging

**DOI:** 10.64898/2026.10.14.26365550

**Authors:** Alexandra Badea, Jimin Xu, Daniel T. Laskowitz, Cristian T. Badea

**Affiliations:** Quantitative Imaging and Analysis Laboratories, Radiology Department, Duke University Medical Center, Durham, NC, USA; Neurology Department, Duke University Medical Center, Durham, NC, USA; Brain Imaging and Analysis Center, Duke University Medical Center, Durham, NC, USA; Biomedical Engineering Department, Duke University, Durham, NC, USA; Department of Biostatistics and Bioinformatics, Duke University School of Medicine, Durham, NC, USA

**Keywords:** APOE ε4, left atrial remodeling, heart–brain axis, brain aging, lateral ventricles, Framingham Heart Study, Alzheimer’s disease, vascular contributions to cognitive impairment and dementia

## Abstract

**INTRODUCTION:** Whether APOE ε4 modifies relationships between subclinical cardiac phenotypes and brain structure is unknown.

**METHODS:** In Framingham Heart Study Offspring participants, we related antecedent echocardiographic remodeling (Examinations 4–6) to repeated brain MRI, and cardiac magnetic resonance phenotypes to regional cortical morphometry, testing cardiac × APOE ε4 interactions with false-discovery-rate (FDR) correction and permutation testing of the regional screen.

**RESULTS:** APOE ε4 modified the association between antecedent left atrial (LA) remodeling and subsequent lateral ventricular expansion (β=−0.00858, P=1.45×10⁻⁴, q=0.0023; N=1,305; 3,787 MRI observations), the only FDR-significant result of 16 longitudinal tests, including with participant-specific MRI-time slopes (q=0.034); it weakened when intracranial volume × MRI time was modeled. Four regional ventricular cardiac × APOE ε4 interactions met within-family FDR criteria, but their number was consistent with chance (permutation P=0.70). Maximum LA volume index showed an APOE ε4-dependent association with left middle-occipital cortical-thickness variability (whole-brain q=0.017; permutation P=0.041) that persisted after ventricular adjustment. Cardiac phenotypes and trajectories showed no FDR-significant APOE ε4-dependent associations with plasma Aβ42/40, p-tau181, GFAP, or NfL, and adjustment for these biomarkers changed the regional interactions by at most 3.2%. Allen Human Brain Atlas data showed cholesterol-biosynthesis enrichment in the right superior frontal cortex, implicated by the LV end-systolic volume interaction, but without convergent pathway evidence.

**DISCUSSION:** APOE ε4 modified atrial–brain relationships, with the strongest evidence linking antecedent atrial remodeling to later ventricular expansion. Replication with prespecified models is needed.

**Highlights:**

- APOE ε4 modified how left atrial remodeling related to later brain ventricle growth.
- It was the only FDR-significant result of 16 tests and persisted with random slopes.
- In ε4 carriers, larger LA volume tracked lower occipital thickness variability.
- This atrial–cortical association was independent of LV mass and ejection fraction.
- Exploratory: HbA1c related to LV end-diastolic volume differently by APOE ε4 status.

## 1 Introduction

Cardiovascular and neurodegenerative processes are increasingly understood as interconnected contributors to late-life cognitive impairment and dementia. Heart failure, atrial fibrillation, coronary disease, and vascular risk factors are associated with cognitive decline and adverse brain outcomes, but clinically overt cardiovascular disease may represent only the later end of a continuum. Subclinical alterations in cardiac output, chamber remodeling, and myocardial function may influence brain health through chronic changes in cerebral perfusion, vascular pulsatility, embolic risk, endothelial function, and blood–brain barrier integrity (Testai et al. 2024). Identifying these relationships before the onset of clinical heart failure, stroke, or dementia may help define modifiable contributors to brain aging (Jefferson 2010; Jefferson et al. 2010).

Population-based imaging studies support associations between cardiac physiology and brain structure even among adults without overt cardiovascular disease. In the Framingham Heart Study, lower cardiac index was associated with smaller total brain volume, greater ventricular enlargement, and slower information processing (Jefferson 2010; Jefferson et al. 2010), and subsequently with incident dementia and Alzheimer disease in participants free of dementia at baseline (Jefferson et al. 2015). Cardiac magnetic resonance studies of left ventricular ejection fraction suggested that both low and high values may be associated with poorer cognitive performance, although continuous associations with global brain measures were limited (Jefferson et al. 2011). Left atrial remodeling may also be relevant to brain health beyond clinically recognized atrial fibrillation. In community-based cohorts, impaired left atrial function has been associated with incident dementia and with MRI evidence of vascular brain injury, including cerebral microbleeds and infarcts, even among participants without prevalent atrial fibrillation or stroke (Wang et al. 2022; Wang et al. 2025). Together, these observations suggest that ventricular pump function, ventricular remodeling, and atrial remodeling capture complementary dimensions of cardiovascular contributions to brain aging.

The apolipoprotein E ε4 (APOE ε4) allele, the strongest common genetic risk factor for late-onset Alzheimer disease, influences lipid and cholesterol transport, neuroinflammation, cerebrovascular function, blood–brain barrier integrity, and cellular energetics (Blumenfeld et al. 2024). APOE ε4 may also modify susceptibility to vascular brain injury; an earlier population study reported greater brain abnormalities when APOE ε4 co-occurred with vascular disease (DeCarli et al. 1999). APOE ε4 is also associated with cardiac phenotype itself. In large British cohorts, carriers had LVEF and LV mass similar to noncarriers but higher myocardial contraction fraction and more favorable strain rates (Topriceanu et al. 2024). A given cardiac measurement may therefore reflect different hemodynamic states in carriers and noncarriers, so APOE ε4 could alter the form of cardiac–brain associations as well as their strength. Most recently, lower cardiac output was associated with smaller brain volumes and faster longitudinal cerebral atrophy predominantly among APOE ε4 carriers (Moore et al. 2026), providing evidence that APOE ε4 modifies heart–brain relationships.

Several questions remain open. First, which cardiac phenotypes carry genotype-dependent associations: in the UK Biobank, cardiac magnetic resonance traits across all four chambers were associated with regional brain morphometry (Zhao et al. 2023), but whether APOE ε4 modifies these phenotype-specific associations has not been examined. Left atrial enlargement may reflect cumulative hemodynamic load and atrial myopathy not captured by ventricular measures. Second, where in the brain associations arise: APOE ε4 studies have relied on global or lobar measures that may obscure localized patterns. Third, whether cardiac remodeling precedes brain change: serial cardiac measurements obtained before brain imaging can establish that order.

We addressed these questions in two complementary Framingham Heart Study Offspring analyses. First, we asked whether left atrial and ventricular remodeling measured across serial echocardiograms was associated with subsequent brain structural change differently by APOE ε4 status. Second, we related cardiac magnetic resonance measures of ventricular function, ventricular remodeling, and left atrial size to regional cortical morphology, with sensitivity analyses for imaging interval, vascular and metabolic risk, atrial fibrillation, familial relatedness, and allele dose, and characterized implicated regions with reference transcriptomic data. The longitudinal analysis establishes measurement order but, like the regional analysis, cannot establish causality.

## Research in Context

1. **Systematic review:** The authors reviewed the literature using PubMed. Population studies have linked cardiac output and left atrial abnormalities to brain structure, vascular brain injury, cognition, and dementia (Jefferson 2010) (Jefferson et al. 2015) (Wang et al. 2022) (Wang et al. 2025), and large imaging studies have mapped cardiac–brain structural associations across many cardiac phenotypes (Zhao et al. 2023). Recent longitudinal work showed that lower cardiac output was associated with faster brain atrophy predominantly in APOE ε4 carriers (Moore et al. 2026). Whether APOE ε4 modifies associations of specific cardiac ventricular and atrial phenotypes, including cardiac remodeling measured before brain MRI, with brain structure had not been examined.
2. **Interpretation:** Among 16 longitudinal heart–brain tests, only the association between antecedent left atrial enlargement and subsequent lateral ventricular expansion differed by APOE ε4 status after correction for multiple comparisons, and it persisted with participant-specific MRI-time slopes. Cross-sectionally, larger left atrial volume was associated with lower left middle-occipital cortical-thickness variability in ε4 carriers (whole-brain q=0.017; permutation P=0.041). Regional cardiac ventricular–cortical interactions were exploratory, and their number was consistent with chance. The associations did not follow a uniform pattern of greater adverse cardiac effects in carriers, suggesting that APOE ε4 may change the form of heart–brain relationships rather than only their strength. The findings are observational and do not establish causality or Alzheimer’s disease specificity.
3. **Future directions:** Replication with prespecified models in independent, more diverse cohorts with repeated cardiac and brain imaging is needed, ideally with atrial rhythm monitoring, cerebral perfusion and vascular-injury measures, and longitudinal Alzheimer’s disease biomarkers and cognition, to test whether the atrial findings generalize and to identify the processes that link cardiac remodeling to brain change in carriers.

## 2 Methods

### 2.1 Study population and design

We analyzed the Framingham Heart Study Offspring Cohort, comprising descendants of the Original Cohort and their spouses (Feinleib et al. 1975), using two complementary observational designs. Controlled-access Framingham data were obtained through dbGaP (phs000007).

The regional imaging arm paired cardiovascular magnetic resonance (CMR) measures with structural brain MRI. The initial imaging overlap included 785 participants; 769 had classified APOE genotype, and the fully adjusted principal regional models included 765 participants. CMR and brain MRI were acquired at separate visits. CMR preceded brain MRI in 782 of 785 participants (99.6%); the mean CMR-to-MRI interval was 2.96 ± 1.26 years (median, 2.87 years; IQR, 2.05–3.72; range, 0.008–7.72 years). Models adjusted for the absolute CMR–MRI interval to account for temporal separation between imaging assessments. Because cardiac and brain imaging were not acquired contemporaneously, associations from this arm were interpreted without assigning temporal direction.

The longitudinal arm related antecedent echocardiographic remodeling across Offspring Examinations 4–6 to repeated subsequent brain MRI. Cardiac trajectories were estimated from serial measurements obtained before the eligible brain MRI observations, thereby establishing measurement order between the cardiac exposure period and subsequent brain follow-up. Model-specific eligibility and complete-case requirements determined sample size in both analytic arms. Participants provided informed consent under applicable Framingham protocols; this secondary analysis used controlled-access data under an approved data-use authorization.

### 2.2 Cardiac phenotypes

CMR-derived principal regional exposures were left ventricular ejection fraction (LVEF), end-systolic volume indexed to body surface area (LVESVi), and indexed LV mass (LVMI). CMR acquisition and computer-aided analysis of left ventricular volumes, mass, and ejection fraction in the Framingham Offspring cohort have been described previously (Chuang et al. 2014). Additional analyses evaluated indexed end-diastolic volume (LVEDVi), left atrial maximum volume index (LAVI), atrial function, and myocardial strain. Ventricular volumes and mass were indexed to body surface area; LVEF was analyzed as a percentage. In the longitudinal arm, echocardiograms at Examinations 4, 5, and 6 provided left atrial dimension, derived LV mass, LV end-diastolic dimension, and derived fractional shortening. Examinations were generally approximately 3–4 years apart. For each measure, a participant-specific ordinary-least-squares slope was fitted against the actual examination dates expressed in years, without assuming uniform spacing. Principal trajectory analyses required a non-missing measurement at all three examinations. The last examination in this exposure window preceded eligible brain MRI observations.

### 2.3 Brain MRI and APOE genotype

Structural MRI and FreeSurfer-derived data provided hemisphere-specific cortical volume, surface area, thickness, and additional morphometric measures. In primary regional volume models, regional volume was divided by intracranial volume (ICV) before standardization; raw regional volume with ICV entered explicitly as a covariate was examined separately. Surface-area models adjusted for total cortical surface area. In addition to mean cortical thickness, we analyzed the regional cortical thickness standard deviation (TKSD, mm) provided in the Framingham FreeSurfer-derived morphometric dataset as a measure of within-region cortical thickness variability.

Longitudinal outcomes included repeated hippocampal volume, total brain volume, lateral ventricular volume, and white matter hyperintensity burden. Eligible participants had at least two post-exposure observations for the outcome being modeled. MRI follow-up time was measured in years since the first eligible MRI. For each longitudinal model, the cardiac trajectory was standardized across participants and the brain outcome across eligible observations; longitudinal coefficients therefore represent change in standardized brain outcome per year.

APOE ε4 carriers had at least one ε4 allele; noncarriers were the reference group. APOE ε4 allele-dose and categorical-genotype analyses were performed as follow-up sensitivity analyses. APOE genotype frequencies were summarized descriptively; genotype categories with very small cell counts were not analyzed separately. APOE ε4 dose analyses were restricted to ε3/ε3, ε3/ε4, and ε4/ε4 genotypes.

### 2.4 Secondary biomarkers and cognition

Secondary analyses considered plasma amyloid-β42/40, phosphorylated tau 181, glial fibrillary acidic protein, and neurofilament light chain. These markers were added to targeted regional cardiac–brain models with matched-complete-case comparisons, were modeled as outcomes of cardiac phenotype × APOE ε4 interactions in the regional cohort, and were separately modeled as later outcomes of antecedent cardiac trajectories. Repeated Mini-Mental State Examination (MMSE)-based scores were used to assess later global cognitive change. Four neuropsychological tests (Logical Memory delayed recall, Trails B, animal fluency, and Digit Span Backward), assessed at the first eligible later examination, were exploratory because repeated detailed testing was less complete. Specifications and correction families are detailed in Supplementary Methods.

### 2.5 Molecular analyses

We obtained adult postmortem microarray gene-expression data from the Allen Institute for Brain Science’s Allen Human Brain Atlas (https://human.brain-map.org/static/download) and mapped them to cortical regions using abagen (Markello et al. 2021). These reference data, rather than participant-level Framingham transcriptomics, were used to characterize genes and biological processes enriched within regions implicated by the regional imaging analyses. This regional reference-atlas enrichment is distinct from testing whether pathway-expression patterns spatially correspond to whole-cortex cardiac × APOE ε4 interaction maps. Participant-level whole-genome sequencing tested pathway-level cardiac effect modification. Separate AHBA analyses tested prespecified pathways and exploratory Gene Ontology/Reactome, MSigDB Hallmark, and brain cell-type sets against the cortical interaction maps, using spatial nulls and analysis-specific FDR correction. Molecular findings were considered descriptive or exploratory and did not establish mediation. Detailed procedures and the reference-atlas provenance are in Supplementary Methods.

### 2.6 Statistical analysis

For each hemisphere-specific brain measure, multivariable ordinary-least-squares regression estimated the cardiac phenotype × APOE ε4 interaction using HC3 robust standard errors. The fully adjusted regional model included age at brain MRI, sex, absolute CMR–MRI interval, body mass index, systolic blood pressure, smoking, diabetes, and the applicable global morphology measure. Continuous variables were standardized within model-specific complete cases. Cortical regions and subcortical structures were assigned to anatomical families by label: 28 Desikan–Killiany and 35 Destrieux regions per hemisphere were assigned to eight anatomical families (seven cortical and one subcortical-nuclei family; hippocampus and amygdala in the medial temporal family) and analyzed for volume, surface area, and thickness, together with seven bilateral subcortical volume measures, yielding 392 brain outcomes per cardiac phenotype and 1,960 tests across five cardiac phenotypes; regions whose labels matched no family were not tested (Supplementary Methods), and Benjamini–Hochberg false-discovery-rate (FDR) correction was applied to interaction *P* values within each of 110 brain metric × anatomical family × cardiac phenotype testing groups, with eligible Desikan–Killiany and Destrieux parcels pooled within each anatomical family. The family structure was finalized after initial analyses. To evaluate the regional screen as a whole, we used a Freedman–Lane permutation test (5,000 permutations): residuals from the fully adjusted model without the cardiac × APOE ε4 term were permuted jointly across all 1,960 regional tests (five cardiac phenotypes × 392 regional outcomes), preserving the APOE ε4 main effect, covariate structure, and correlation among regions, and the complete analysis, including within-family FDR correction, was repeated for each permutation. As a post hoc focused sensitivity analysis, the minimum-P permutation test was repeated in a non-redundant subset comprising the 28 eligible Desikan–Killiany cortical regions per hemisphere and the hippocampal, amygdalar, and subcortical nuclear volumes, for the three principal ventricular phenotypes and all three morphometric types (546 tests). Atrial phenotypes were analyzed in a separate screen in which Benjamini–Hochberg correction was applied across all 1,086 label-selected FreeSurfer morphometric outcomes (Desikan–Killiany and Destrieux volume, surface area, thickness, thickness variability, and mean curvature, plus six additional label-matched variables; Supplementary Methods) within each atrial phenotype; the LAVI screen was also evaluated with a 5,000-permutation Freedman–Lane minimum-P test preserving outcome-specific complete cases. Findings with within-family q<0.05 were treated as exploratory and underwent targeted APOE-stratified, imaging-interval, vascular/metabolic, atrial-fibrillation, pedigree-family clustering, and genotype sensitivity analyses. Because CMR and brain MRI were acquired at separate visits, these regional analyses were interpreted as associations without temporal direction.

For the longitudinal analysis, each combination of four antecedent cardiac trajectories and four subsequent brain outcomes was evaluated using a participant-specific random-intercept mixed-effects model. Models included MRI time, standardized cardiac trajectory, APOE ε4 carrier status, all corresponding two-way interactions, and the cardiac trajectory × MRI time × APOE ε4 interaction, with adjustment for age at Examination 6 and sex. The three-way interaction tested whether the association between antecedent cardiac remodeling and subsequent brain-change rate differed according to APOE ε4 status. Eligible participants had measurements at all three cardiac Examinations 4–6 and at least two MRI observations after the final cardiac assessment. The median interval from the final cardiac assessment to first eligible MRI was 3.47 years (IQR, 2.83–4.28), followed by a median MRI follow-up of 12.45 years (IQR, 6.84–13.94). A temporal sensitivity analysis required an interval of at least two years between the final cardiac assessment and first MRI. Cardiac trajectories and brain outcomes were standardized within each eligible model sample; longitudinal coefficients therefore represent differences in annual change in standardized brain outcome. Benjamini–Hochberg FDR correction was applied to the 16 trajectory–outcome three-way interaction tests separately within each temporal subset. All 16 models were also refitted with participant-specific random intercepts and MRI-time slopes, together with age × MRI time and sex × MRI time terms, with FDR correction across the 16 tests. Genotype-stratified slopes, vascular and metabolic adjustment (including Examination 4 antihypertensive treatment), APOE ε4 genotype analyses, and generalized estimating equations were targeted follow-up analyses; GEE results were subjected to their own multiplicity correction. Longitudinal genotype sensitivity analyses were restricted to participants with ε3/ε3, ε3/ε4, or ε4/ε4 genotypes, with ε3/ε3 as the reference group. Genotype-specific LA remodeling × MRI time effects were estimated with the same specification as the primary carrier model, and a 2-df Wald test assessed overall genotype modification.

Targeted sensitivity analyses evaluated longitudinal model specification and the definition of antecedent atrial remodeling. These analyses added age at Examination 6 × MRI time and sex × MRI time interactions, incorporated a subject-specific random MRI-time slope, and further adjusted for baseline LA dimension and baseline LA dimension × MRI time. An alternative remodeling measure was calculated as annualized change in LA dimension directly from Examination 4 to Examination 6. Head-size sensitivity analyses were performed in the ICV-complete subset using otherwise matched models without ICV terms, with ICV as a covariate, and with ICV and ICV × MRI time, allowing effects of sample restriction to be distinguished from those of additional ICV adjustment. Native-unit interpretation of the LA remodeling interaction was obtained by back-transforming the standardized estimate using the observed standard deviations of LA remodeling and lateral ventricular volume. Detailed specifications are provided in the Supplementary Methods.

To evaluate potential selection into the repeated-MRI longitudinal cohort, we defined a pre-MRI at-risk cohort of participants with valid three-examination LA trajectories and classified APOE genotype. Inclusion in the primary longitudinal analysis was defined as having at least two eligible subsequent lateral-ventricular MRI observations. Descriptive MRI availability was summarized overall and according to APOE ε4 carrier status and tertiles of antecedent LA remodeling. The probability of inclusion was modeled using standardized LA remodeling, APOE ε4 carrier status, their interaction, age at Examination 6, sex, Examination 4 body mass index, systolic blood pressure, diabetes, and smoking, and baseline LA dimension as predictors. Stabilized inverse-probability-of-selection weights were derived from the fitted probabilities and truncated at the 1st and 99th percentiles. The longitudinal LA remodeling × MRI time × APOE ε4 association was then re-estimated using weighted generalized estimating equations and compared with an unweighted GEE fitted in the same complete-case sample.

All tests were two-sided and used complete cases. Participant and observation counts, nominal *P* values, FDR-adjusted q values, and applicable correction families are reported with the corresponding results. Temporal ordering in the longitudinal analyses was interpreted as supporting temporality but not causality.

### Ethics approval and consent to participate

The Framingham Heart Study protocols were reviewed by the Institutional Review Board of Boston University Medical Center, and participants provided written informed consent. The Duke University Health System Institutional Review Board determined that this analysis of de-identified data did not constitute human subjects research. Data were accessed through dbGaP under authorized access (phs000007).

### Code-availability statement

Analysis code, including scripts that regenerate all figures and tables and verify them against the reported values, is available at https://github.com/AD-Decode/atrial_brain.

## 3 Results

### 3.1 Study samples and analytic cohorts

The study included complementary regional imaging and longitudinal analytic cohorts (Figure 1).

**Figure 1.**
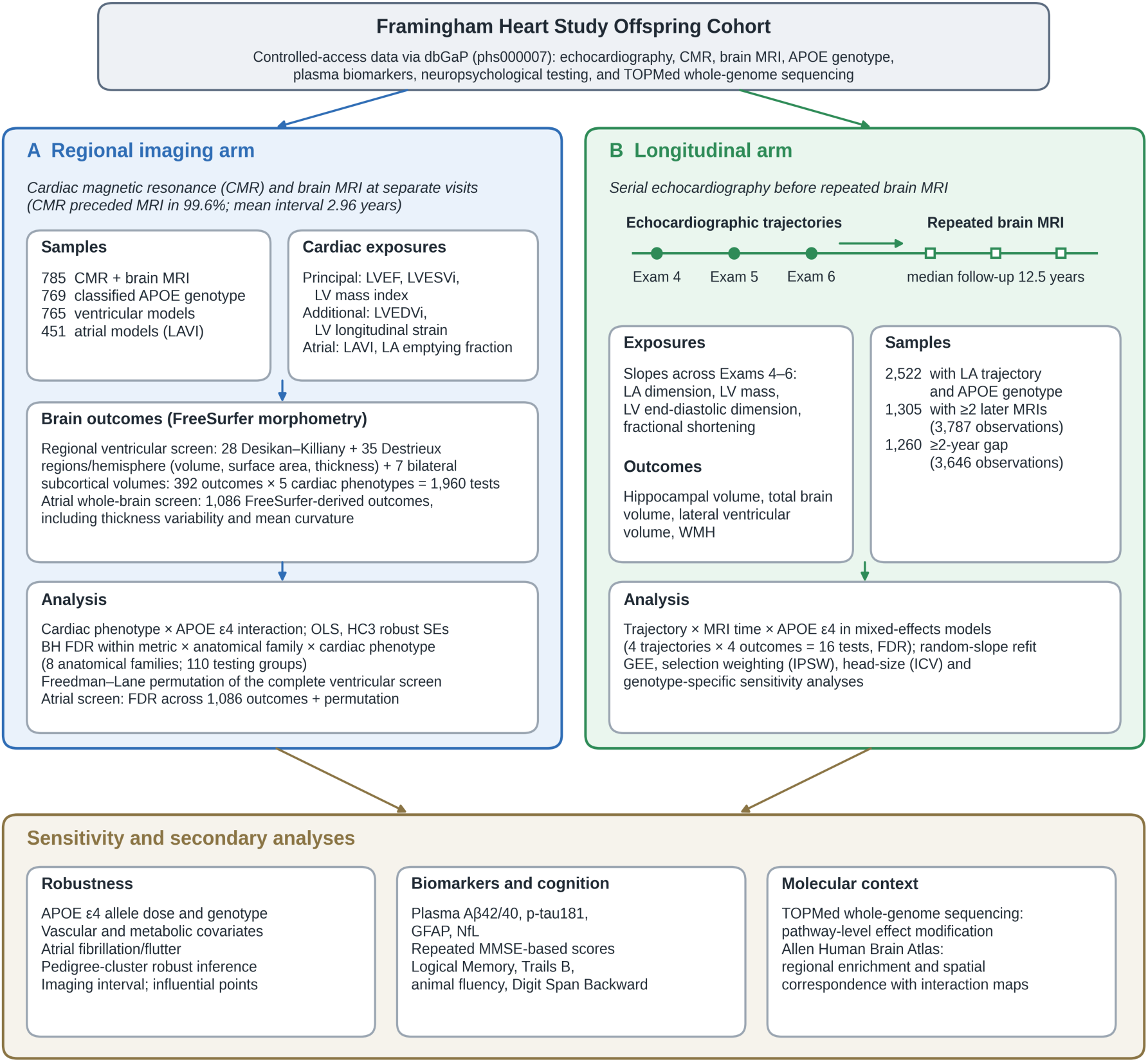
Study design and analytic framework. The Framingham Offspring cohort was analyzed in two complementary arms. (A) The regional imaging arm included 785 participants with cardiac magnetic resonance (CMR) and brain MRI acquired at separate visits (765 in ventricular and 451 in atrial models). The regional ventricular screen comprised 28 Desikan–Killiany and 35 Destrieux regions per hemisphere (volume, surface area, thickness) and seven bilateral subcortical volumes, giving 392 outcomes × 5 cardiac phenotypes = 1,960 tests, with Benjamini–Hochberg FDR correction within 110 metric × anatomical family × cardiac phenotype groups and a Freedman–Lane permutation of the complete screen; the atrial whole-brain screen comprised 1,086 FreeSurfer-derived outcomes. (B) The longitudinal arm estimated left atrial and ventricular trajectories from echocardiography at Offspring Examinations 4–6 and related them to subsequent repeated brain MRI (1,305 participants, 3,787 observations; 1,260 participants and 3,646 observations with at least 2 years between the final cardiac assessment and first eligible MRI). Secondary analyses addressed APOE ε4 allele dose and genotype, vascular and metabolic covariates, atrial fibrillation/flutter, familial clustering, plasma Alzheimer disease–related biomarkers, cognition, and molecular context.

The regional imaging cohort comprised 785 Framingham Heart Study Offspring participants with both cardiac magnetic resonance (CMR) and brain MRI data; APOE genotype was classified in 769 participants, including 596 APOE ε4 noncarriers and 173 carriers (Table 1). Among participants with classified APOE genotype, mean age at CMR was 61.77 years (SD, 8.22), 448 (58.3%) were women, mean body mass index was 27.82 kg/m² (SD, 4.77), 73 (9.5%) had diabetes, and 49 (6.4%) were current smokers. CMR and brain MRI were acquired at separate visits, with a mean absolute interval of 2.96 years (SD, 1.25). The primary ventricular cardiac × APOE ε4 models included 765 participants after model-specific complete-case exclusions. Maximum left atrial volume index was available in a smaller subset, yielding 451 participants for the primary atrial interaction model. Cardiac characteristics were generally similar between APOE ε4 carriers and noncarriers, although maximum LAVI was modestly lower among carriers (Table 1).

**Table 1.** Characteristics of the cross-sectional and longitudinal analytic cohorts. Panel A summarizes the cross-sectional CMR–brain MRI cohort; Panel B summarizes the primary prospective longitudinal LA-dimension structural-MRI cohort.

| **Panel A. Cross-sectional CMR–brain MRI cohort** |  |  |  |  |
| --- | --- | --- | --- | --- |
| Characteristic | APOE-classified sample<br>(N = 769) | APOE $\epsilon 4$ noncarriers<br>(n = 596) | APOE $\epsilon 4$ carriers<br>(n = 173) | P value |
| **Demographic and clinical characteristics** |  |  |  |  |
| Age at CMR, years | 61.77 ± 8.22 | 61.87 ± 8.29 | 61.40 ± 8.00 | 0.503 |
| Female sex, n (%) | 448 (58.3%) | 347 (58.2%) | 101 (58.4%) | 1.0 |
| BMI, kg/m<sup>2</sup> | 27.82 ± 4.77 | 27.76 ± 4.79 | 28.00 ± 4.69 | 0.556 |
| Systolic blood pressure, mmHg | 126.28 ± 16.29 | 126.38 ± 16.25 | 125.97 ± 16.46 | 0.772 |
| Diabetes, n (%) | 73 (9.5%) | 56 (9.4%) | 17 (9.8%) | 0.937 |
| Current smoking, n (%) | 49 (6.4%) | 40 (6.7%) | 9 (5.2%) | 0.59 |
| Absolute CMR–brain MRI interval, years | 2.96 ± 1.25 | 2.97 ± 1.23 | 2.94 ± 1.31 | 0.799 |
| **Cardiac imaging characteristics** |  |  |  |  |
| LVEF, % | 60.40 ± 5.60 | 60.45 ± 5.65 | 60.22 ± 5.43 | 0.626 |
| LVEDVi, mL/m<sup>2</sup> | 73.82 ± 13.40 | 74.19 ± 13.51 | 72.55 ± 12.97 | 0.149 |
| LVESVi, mL/m<sup>2</sup> | 29.44 ± 7.88 | 29.56 ± 8.08 | 29.01 ± 7.16 | 0.382 |
| LV mass index, g/m<sup>2</sup> | 45.91 ± 8.86 | 46.14 ± 8.92 | 45.14 ± 8.63 | 0.187 |
| LAVI maximum, mL/m<sup>2</sup> | 28.44 ± 8.01 | 28.80 ± 8.19 | 27.13 ± 7.22 | 0.045 |
| LA total emptying fraction, % | 49.02 ± 6.67 | 48.96 ± 6.63 | 49.25 ± 6.82 | 0.695 |
| **APOE genotype, n (%)** |  |  |  |  |
| $\epsilon 2/\epsilon 2$ | $\leq 5$ ($\leq 0.7\%$) | $\leq 5$ ($\leq 0.8\%$) | 0 (0.0%) | |
| $\epsilon 2/\epsilon 3$ | 107 (13.9%) | 107 (18.0%) | 0 (0.0%) | |
| $\epsilon 2/\epsilon 4$ | 12 (1.6%) | 0 (0.0%) | 12 (6.9%) | |
| $\epsilon 3/\epsilon 3$ | 486 (63.2%) | 486 (81.5%) | 0 (0.0%) | |
| $\epsilon 3/\epsilon 4$ | 148 (19.2%) | 0 (0.0%) | 148 (85.5%) | |
| $\epsilon 4/\epsilon 4$ | 13 (1.7%) | 0 (0.0%) | 13 (7.5%) | |
| **Panel B. Longitudinal cardiac-remodeling–structural brain MRI cohort** |  |  |  |  |
| Characteristic | APOE-classified sample<br>(N = 1,305) | APOE $\epsilon 4$ noncarriers<br>(n = 1,002) | APOE $\epsilon 4$ carriers<br>(n = 303) | P value |
| **Longitudinal imaging characteristics** |  |  |  |  |
| MRI observations, n | 3787 | 2917 | 870 |  |
| MRI observations per participant, median (IQR) | 3 (2–3) | 3 (2–3) | 3 (2–3) |  |
| MRI follow-up, years, median (IQR) | 12.45 (6.84–13.94) | 12.46 (6.87–13.95) | 12.40 (6.78–13.85) |  |
| Last cardiac examination to first brain MRI, years, median (IQR) | 3.47 (2.83–4.28) | 3.46 (2.83–4.28) | 3.49 (2.85–4.30) |  |
| **Participant characteristics** |  |  |  |  |
| Age at Examination 6, years | 56.64 ± 9.13 | 56.80 ± 9.14 | 56.12 ± 9.10 | 0.255 |
| Female sex, n (%) | 685 (52.5%) | 525 (52.4%) | 160 (52.8%) | 0.948 |
| LA dimension trajectory, cm/year | 0.0355 ± 0.0604 | 0.0361 ± 0.0607 | 0.0335 ± 0.0596 | 0.511 |

The longitudinal arm used serial echocardiographic measurements from Offspring Examinations 4–6 to estimate antecedent trajectories of left atrial dimension, left ventricular mass, left ventricular end-diastolic dimension, and fractional shortening. In the principal prospective LA-dimension–lateral-ventricular analysis, 1,305 participants contributed 3,787 subsequent MRI observations (Table 1). Participants had a median of 3 MRI observations (IQR, 2–3), with a median MRI follow-up of 12.45 years (IQR, 6.84–13.94). The median interval between the final cardiac assessment and first eligible brain MRI was 3.47 years (IQR, 2.83–4.28). A temporally more restrictive analysis requiring an interval of at least 2 years included 1,260 participants contributing 3,646 MRI observations. Sample sizes for other longitudinal models varied according to availability of the relevant cardiac trajectory, brain outcome, APOE genotype, and covariates.

### 3.2 Antecedent left atrial remodeling is associated with subsequent ventricular change in an APOE ε4-dependent manner

Antecedent left atrial (LA) remodeling was associated with the subsequent rate of lateral ventricular expansion in an APOE ε4-dependent manner (Figure 2; Table S1). In the principal prospective mixed-effects model, the LA dimension trajectory × MRI time × APOE ε4 interaction was β=−0.00858 (95% CI, −0.01300 to −0.00415; *P*=1.45×10⁻⁴; FDR q=0.00231; N=1,305; 3,787 MRI observations; Table S2). Requiring at least a 2-year interval between the final cardiac assessment and first eligible brain MRI yielded a similar estimate (β=−0.00813, 95% CI, −0.01270 to −0.00356; *P*=4.84×10⁻⁴; FDR q=0.00774; N=1,260), supporting temporal robustness of the association.

**Figure 2.**
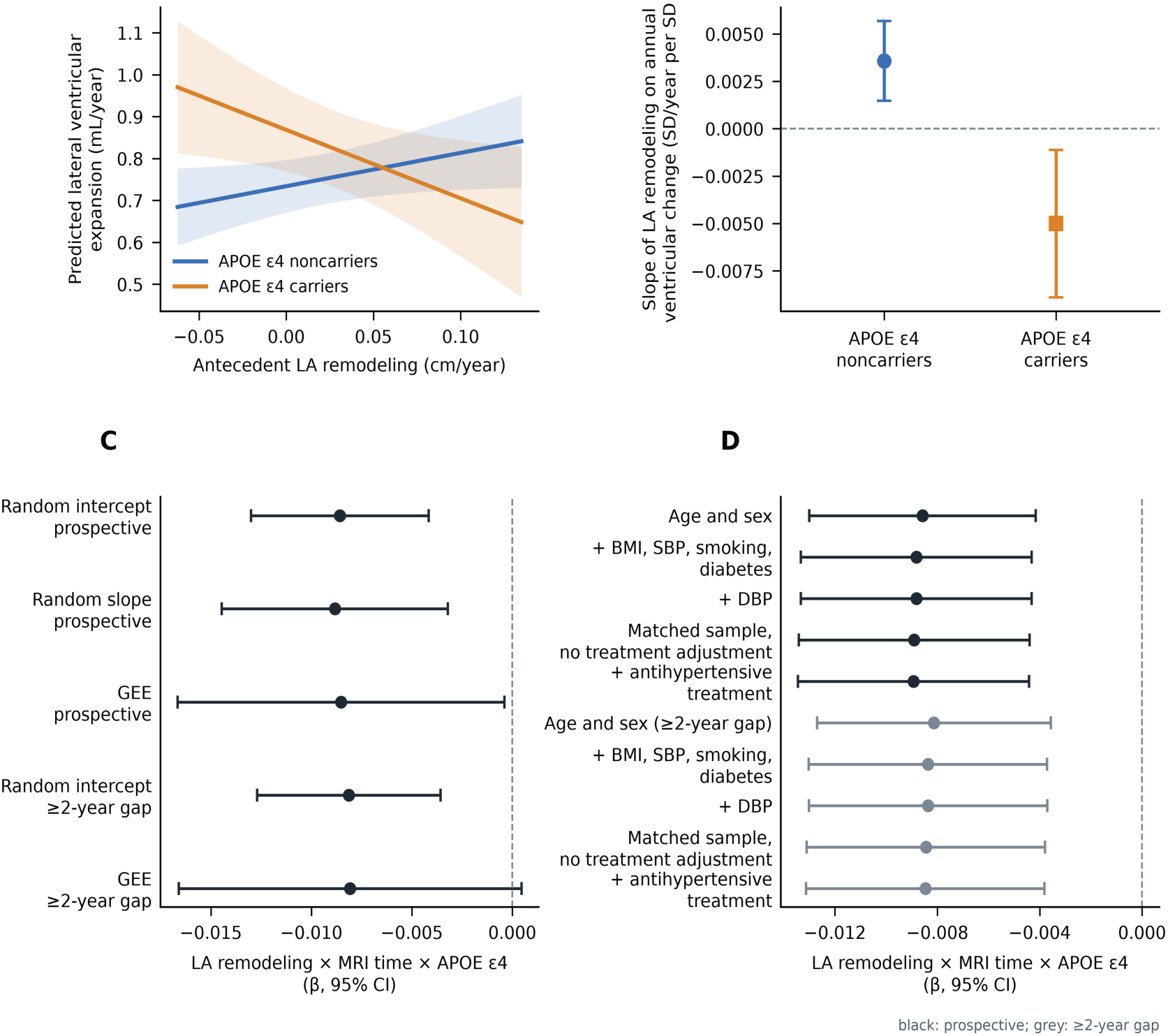
Antecedent left atrial remodeling, APOE ε4, and subsequent lateral ventricular change. (A) Predicted annual change in lateral ventricular volume across antecedent LA remodeling rates for APOE ε4 carriers and noncarriers, from the expanded longitudinal model (age × MRI time, sex × MRI time, baseline LA dimension × MRI time, and participant-specific random MRI-time slopes; interaction β=−0.00880, P=0.0022); shaded bands are 95% confidence intervals (CIs). Each 0.05 cm/year greater LA enlargement corresponded to a −0.121 mL/year shift in the carrier–noncarrier difference in ventricular expansion rate (95% CI, −0.199 to −0.044); predicted expansion remained positive in both groups. (B) APOE ε4-stratified slopes of LA remodeling on annual ventricular change from the principal random-intercept model, in standardized units: noncarriers, β=0.0036 (P=8.8×10⁻⁴; robust GEE P=0.012); carriers, β=−0.0050 (P=0.012; robust GEE P=0.20). (C) The LA remodeling × MRI time × APOE ε4 interaction from the random-intercept, random-slope, and generalized estimating equation (GEE) models in the prospective sample and from the random-intercept and GEE models in the ≥2-year-gap sample. \(D) The interaction after adjustment for Examination 4 vascular and metabolic covariates and, in the matched sample with treatment data (N=1,268), for antihypertensive treatment; black, prospective sample; grey, ≥2-year-gap sample. In the principal model, β=−0.00858 (95% CI, −0.01300 to −0.00415; P=1.45×10⁻⁴; FDR q=0.0023; N=1,305); with random MRI-time slopes across all 16 longitudinal models, it was the only FDR-significant interaction (P=0.0021; q=0.034).

Stratified slopes showed that faster antecedent LA enlargement was associated with faster subsequent lateral ventricular expansion in APOE ε4 noncarriers (β=0.0036; 95% CI, 0.0015 to 0.0057; *P*=8.8×10⁻⁴; robust GEE *P*=0.012). In carriers, the slope was negative in the mixed model (β=−0.0050; 95% CI, −0.0089 to −0.0011; *P*=0.012) but not distinguishable from zero with robust standard errors (*P*=0.20). The interaction therefore reflects the presence of the expected association in noncarriers and its absence in carriers. Predicted native-unit trajectories showed ventricular expansion across the illustrated range of LA remodeling in both groups (Figure 2A), so the interaction does not imply ventricular contraction among carriers.

Additional adjustment for Examination 4 vascular covariates (body mass index, systolic blood pressure, smoking, and diabetes) yielded a similar prospective interaction estimate (β=−0.00882, 95% CI, −0.01333 to −0.00431; *P*=1.25×10⁻⁴; N=1,269; 3,685 MRI observations), with essentially unchanged results after additional adjustment for diastolic blood pressure (β=−0.00882, 95% CI, −0.01333 to −0.00431; *P*=1.26×10⁻⁴; Table S3). The interaction was also unchanged after additional adjustment for antihypertensive treatment at Examination 4: in the matched sample with treatment data (N=1,268), β=−0.00890 without and β=−0.00893 with treatment adjustment (P=1.05×10⁻⁴; Table S3), a change of less than 0.3% in absolute effect size. Generalized estimating equations yielded a similar point estimate (β=−0.00852) but greater uncertainty (*P*=0.0399), and the interaction did not survive the corresponding FDR correction (q=0.368; Table S1).

In sensitivity analyses restricted to ε3/ε3, ε3/ε4, and ε4/ε4 participants, the association between antecedent LA remodeling and subsequent lateral ventricular change differed across genotype groups (2-df Wald χ²=10.50, *P*=0.0053), but the pattern was not monotonic in ε4 allele count (Table S1). The LA remodeling × MRI time estimate was +0.00211 in ε3/ε3 participants (n=823; *P*=0.070), −0.00552 in ε3/ε4 participants (n=256; *P*=0.0091), and +0.00622 in the small ε4/ε4 group (n=23; *P*=0.442). These analyses support genotype heterogeneity but not a longitudinal ε4 allele-dose response.

Targeted sensitivity analyses further evaluated longitudinal model specification and the definition of antecedent atrial remodeling (Table S3). The interaction was preserved after inclusion of age × MRI time and sex × MRI time terms and a participant-specific random MRI-time slope (β=−0.00883, 95% CI, −0.01445 to −0.00320; P=0.00210; q=0.034 when all 16 longitudinal models were refitted with this specification, in which it was the only FDR-significant result; Table S2), and remained similar after additional adjustment for baseline LA dimension and baseline LA dimension × MRI time (β=−0.00880, 95% CI, −0.01442 to −0.00317; **P=0.00223**). Defining atrial remodeling instead as annualized LA dimensional change directly from Examination 4 to Examination 6 yielded a consistent association (β=−0.00759, 95% CI, −0.01305 to −0.00212; P=0.00650; N=1,356; 3,924 MRI observations).

Head-size sensitivity analyses showed attenuation that reflected both restriction to the ICV-complete sample and additional longitudinal ICV adjustment. In the ICV-complete subset (N=795; 2,405 MRI observations), the matched random-intercept model without ICV terms yielded an interaction estimate of β=−0.00451 (95% CI, −0.00813 to −0.00090; P=0.0144), smaller in magnitude than the principal estimate in the full sample (β=−0.00858; N=1,305). Adjustment for ICV alone produced essentially no further change (β=−0.00449, 95% CI, −0.00811 to −0.00088; P=0.0148). When ICV × MRI time was additionally included, the interaction was further attenuated to β=−0.00334 (95% CI, −0.00680 to 0.00011; P=0.0581). Thus, much of the attenuation was already present after restriction to participants with available ICV, while modeling longitudinal ICV effects produced additional attenuation and reduced precision.

For interpretation in native units, using the expanded random-slope model, each 0.05 cm/year (0.5 mm/year) greater antecedent rate of LA enlargement corresponded to a −0.121 mL/year shift in the APOE ε4 carrier–noncarrier difference in subsequent lateral ventricular expansion rate (95% CI, −0.199 to −0.044; *P*=0.0022; Figure 2A).

Because the longitudinal analysis required continued participation through repeated brain MRI, we additionally examined potential selection bias. Among 2,522 participants with valid three-examination LA trajectories and classified APOE genotype, 1,305 (51.7%) had at least two eligible subsequent MRI examinations, 339 (13.4%) had one eligible MRI, and 878 (34.8%) had no eligible MRI. Descriptively, inclusion rates were similar across APOE ε4 status and tertiles of antecedent LA remodeling, ranging from 51.4% to 60.6% among ε4 carriers and from 48.7% to 53.2% among noncarriers (Table S4). In the multivariable selection model, the LA remodeling × APOE ε4 interaction was not associated with probability of inclusion in the repeated-MRI cohort (OR=1.065, 95% CI 0.880–1.288; P=0.519; Table S4). Inverse-probability-of-selection weighting yielded a similar LA remodeling × MRI time × APOE ε4 interaction (β=−0.00839, 95% CI −0.01630 to −0.00049; P=0.0373; N=1,269; 3,685 MRI observations), compared with β=−0.00869 (95% CI −0.01683 to −0.00056; P=0.0361) in the unweighted GEE restricted to the same complete-case sample (Table S4).

### 3.3 Antecedent cardiac remodeling, later plasma biomarkers, and cognition

Antecedent cardiac trajectories showed no APOE ε4-dependent associations with later plasma Aβ42/40, p-tau181, GFAP, or NfL after correction across the 16 trajectory × biomarker tests (minimum q=0.740; Table S5). Across 16 analyses of four later cognitive tests, no cardiac trajectory × APOE ε4 interaction survived correction (minimum q=0.179; Table S5, Panel A); the LV end-diastolic dimension × APOE ε4 interaction for Digit Span Backward was nominally significant (*P*=0.0112). In repeated MMSE assessments, the LV mass trajectory × cognitive time × APOE ε4 interaction met the four-test FDR criterion (β=−0.00786, 95% CI, −0.0136 to −0.0021; *P*=0.0073; q=0.029; N=1,205; 6,409 observations; Table S5, Panel B). The LA dimension trajectory, which carried the principal structural finding, showed no corresponding MMSE interaction (*P*=0.53).

### 3.4 APOE ε4 modifies regional ventricular cardiac–brain associations

In a complementary regional CMR–brain MRI analysis, four ventricular cardiac phenotype × APOE ε4 interactions met within-family q<0.05 after FDR correction within their respective brain metric × anatomical family × cardiac phenotype testing groups (Figure 3; Table 2; complete results for all 1,960 tests in Data S1).

**Figure 3.**
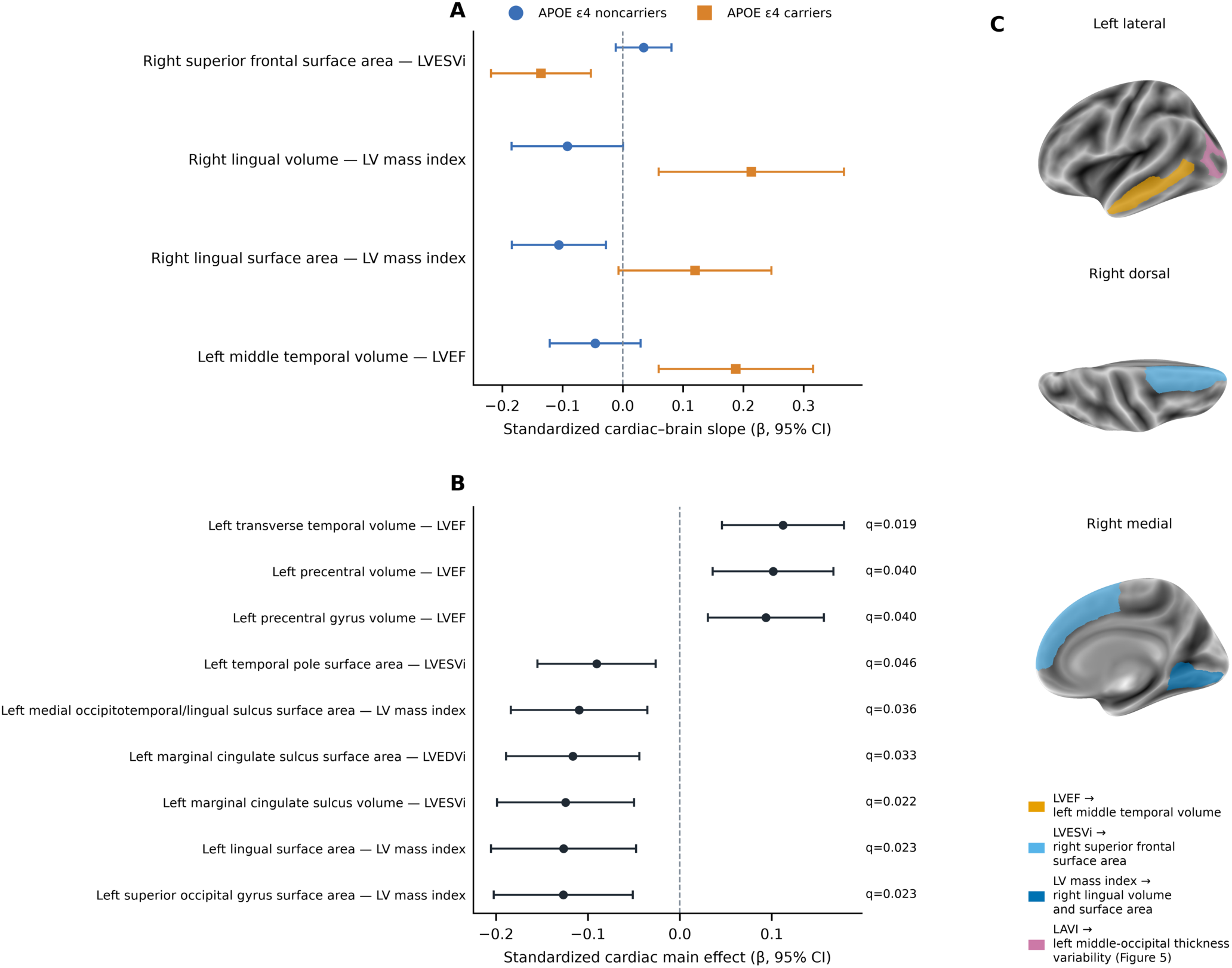
Regional cardiac phenotype × APOE ε4 associations with cortical morphology (exploratory). (A) APOE ε4-stratified standardized cardiac–brain slopes for the four ventricular interactions with within-family q<0.05 (Benjamini–Hochberg correction within brain metric × anatomical family × cardiac phenotype groups); carrier slopes and 95% CIs were computed from the interaction model with its robust covariance. Inference on effect modification is based on the interaction term. In a Freedman–Lane permutation test of all 1,960 regional interaction tests, the number of within-family discoveries was consistent with chance (permutation P=0.70). (B) Regional cardiac main effects with within-family q<0.05 in vascular-adjusted models (N=781). (C) Localization on the inflated fsaverage surface (left lateral, right dorsal, and right medial views): left middle temporal gyrus (LVEF; volume), right superior frontal gyrus (LVESVi; surface area), right lingual gyrus (LV mass index; volume and surface area), and left middle occipital gyrus (LAVI; cortical-thickness variability; Figure 5). Points are standardized coefficients and lines 95% CIs. Interaction estimates are in Table 2, main effects in Table S6, and carrier slopes in Table 2.

**Table 2.** APOE ε4 modification of ventricular cardiac–brain structural associations.

| Cardiac phenotype | Brain phenotype | N | $\beta$, noncarriers / carriers | $\beta$, interaction | 95% CI | P | FDR q |
| --- | --- | --- | --- | --- | --- | --- | --- |
| LVEF | Left middle temporal volume | 765 | −0.046 / 0.188 | 0.233 | 0.087 to 0.380 | 0.00176 | 0.0352 |
| LVESVi | Right superior frontal surface area | 765 | 0.035 / −0.136 | −0.170 | −0.260 to −0.081 | $1.97 \times 10^{-4}$ | 0.00433 |
| LV mass index | Right lingual volume | 765 | −0.092 / 0.213 | 0.305 | 0.144 to 0.467 | $2.16 \times 10^{-4}$ | 0.00604 |
| LV mass index | Right lingual surface area | 765 | −0.106 / 0.120 | 0.226 | 0.095 to 0.357 | $7.09 \times 10^{-4}$ | 0.0198 |

$\beta$ , noncarriers, is the standardized cardiac slope among APOE $\epsilon 4$ noncarriers; $\beta$ , interaction, is the cardiac phenotype × APOE $\epsilon 4$ interaction coefficient and represents the difference in slope for carriers relative to noncarriers. Models were adjusted for age, sex, absolute CMR–brain MRI interval, body mass index, systolic blood pressure, current smoking, diabetes history, and morphology-specific global measures; HC3 robust standard errors were used. FDR q values were calculated separately within each brain metric × anatomical family × cardiac phenotype testing group. APOE, apolipoprotein E; CI, confidence interval; CMR, cardiac magnetic resonance; FDR, false discovery rate; LVEF, left ventricular ejection fraction; LVESVi, left ventricular end-systolic volume indexed to body surface area; LV mass index, left ventricular mass indexed to body surface area. Carrier slopes ( $\beta$ noncarrier + $\beta$ interaction) are shown after the slash; their 95% CIs, computed with the robust covariance of the two coefficients, are in Supplementary Data S2.

In the permutation analysis, the number of within-family FDR discoveries did not exceed that expected under the null hypothesis of no cardiac × APOE ε4 interaction (observed, 4; null median, 6; permutation P=0.70), and at least one within-family discovery occurred in 95% of permutations. The smallest observed interaction P value was also consistent with chance (minimum-P permutation P=0.28). In the post hoc focused subset of 546 non-redundant tests, the strongest interaction did not reach familywise significance (minimum-P permutation P=0.11); an earlier cortical-only subset and metric-specific tests are reported in the Supplementary Methods. The regional ventricular findings are therefore considered exploratory and hypothesis-generating. These analyses assessed regional associations between cardiac and brain structure without establishing temporal direction. Covariate-adjusted regional cardiac main effects meeting the corresponding FDR criterion are shown separately in Figure 3B and Table S6.

Higher LV end-systolic volume index (LVESVi) was differentially associated with right superior frontal surface area according to APOE ε4 status (interaction β=−0.170, 95% CI, −0.260 to −0.081; *P*=1.97×10⁻⁴; q=0.0043; N=765). The association was positive among APOE ε4 noncarriers (β=0.035) but negative among carriers (β=−0.136). LV mass index showed two APOE ε4-dependent associations involving the right lingual cortex. For right lingual volume, the interaction was β=0.305 (95% CI, 0.144 to 0.467; *P*=2.16×10⁻⁴; q=0.0060; N=765), with a negative association among noncarriers (β=−0.092) and a positive association among carriers (β=0.214). For right lingual surface area, the interaction was β=0.226 (95% CI, 0.095 to 0.357; *P*=7.09×10⁻⁴; q=0.0198; N=765), with β=−0.106 among noncarriers and β=0.120 among carriers.

LVEF was differentially associated with left middle temporal volume according to APOE ε4 status (interaction β=0.233, 95% CI, 0.087 to 0.380; *P*=0.00176; q=0.0352; N=765). The association was weakly negative among noncarriers (β=−0.046) and positive among carriers (β=0.188). Inference regarding APOE ε4 effect modification was based on the cardiac phenotype × APOE ε4 interaction terms rather than on the statistical significance of genotype-stratified slopes. Temporal robustness of these regional associations was examined subsequently.

### 3.5 Temporal and morphometric sensitivity analyses

Temporal sensitivity differed across the four primary regional cardiac × APOE ε4 interactions (Figure 4). Using fixed full-cohort standardization, the LVESVi × APOE ε4 association with right superior frontal surface area was preserved after restricting the CMR–brain MRI interval to ≤3 years (β=−0.246, 95% CI, −0.360 to −0.131; *P*=2.65×10⁻⁵; N=418) and ≤2 years (β=−0.296, 95% CI, −0.480 to −0.113; *P*=0.00157; N=185), compared with β=−0.170 (95% CI, −0.260 to −0.081; *P*=1.97×10⁻⁴; N=765) in the full cohort. The LVEF × APOE ε4 association with left middle temporal volume remained positive but became less precise with interval restriction (≤3 years: β=0.165, 95% CI, −0.022 to 0.351; *P*=0.0837; ≤2 years: β=0.331, 95% CI, −0.007 to 0.669; *P*=0.0552), compared with β=0.233 (95% CI, 0.087 to 0.380; *P*=0.00176) in the full cohort. In contrast, the LV mass index × APOE ε4 associations with right lingual volume (full cohort: β=0.305; ≤3 years: β=0.227; ≤2 years: β=−0.032) and surface area (full cohort: β=0.226; ≤3 years: β=0.195; ≤2 years: β=−0.021) attenuated with increasingly stringent interval restriction. Thus, temporal robustness was heterogeneous across the four regional findings, with the superior frontal association preserved under both interval restrictions, the middle temporal association retaining its direction with reduced precision, and the lingual associations attenuating toward the null.

**Figure 4.**
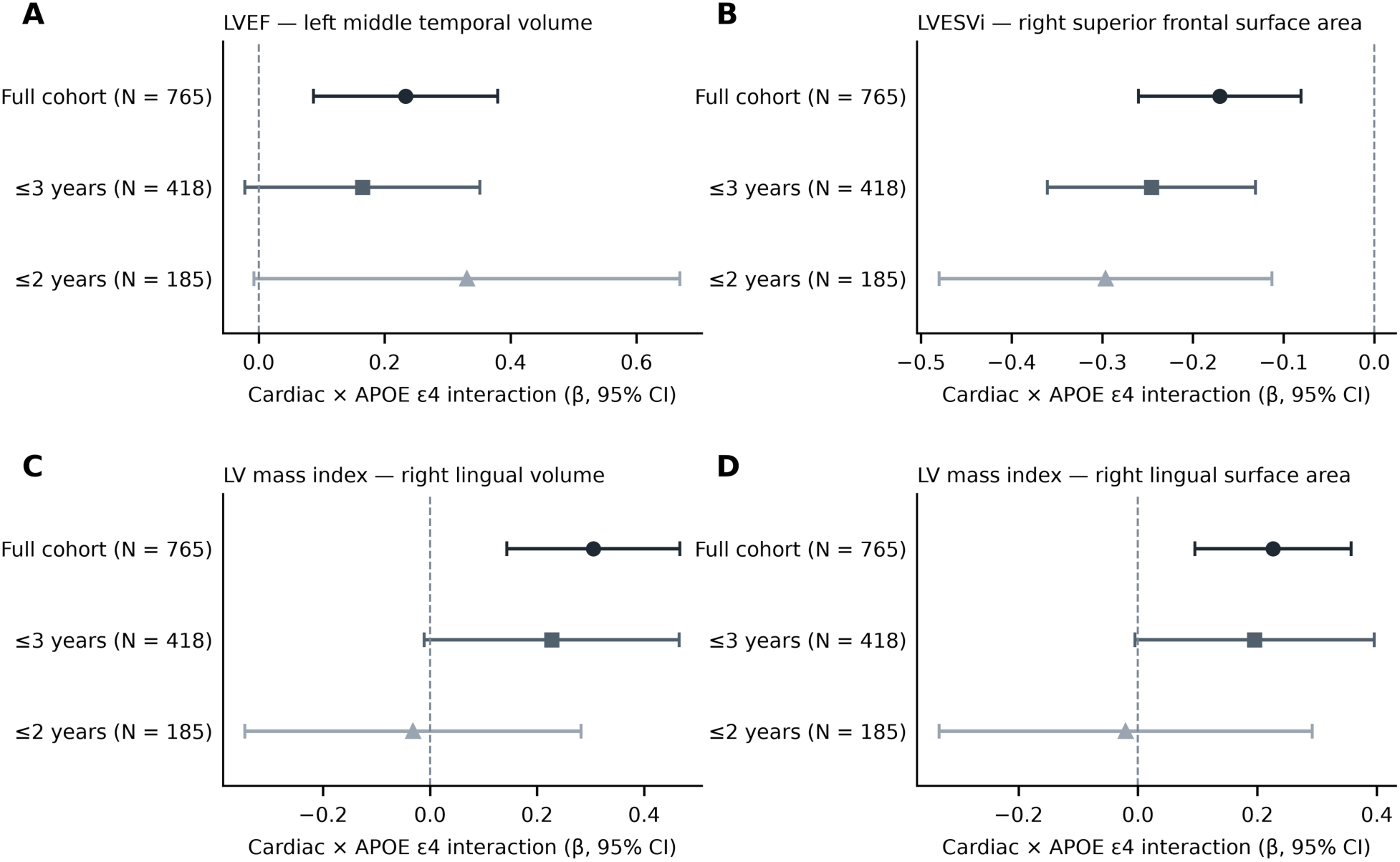
Sensitivity of the regional ventricular interactions to the CMR–brain MRI interval. Cardiac phenotype × APOE ε4 interactions in the full cohort (N=765) and after restricting the CMR–brain MRI interval to ≤3 years (N=418) and ≤2 years (N=185), for (A) LVEF–left middle temporal volume, (B) LVESVi–right superior frontal surface area, (C) LV mass index–right lingual volume, and (D) LV mass index–right lingual surface area. Means and standard deviations of all continuous variables were taken from each full-cohort model and held fixed in the restricted samples, so that coefficients share one scale. Points are interaction coefficients and lines 95% CIs.

The primary ventricular interactions were also evaluated using morphology-specific alternative specifications. Models using raw regional volumes with intracranial volume entered explicitly as a covariate were broadly consistent with the primary ICV-normalized analyses, and surface-area interactions were similar with and without adjustment for total cortical surface area; all alternative estimates retained their direction and excluded zero (Figure S1). Additional metabolic sensitivity analyses showed little change in the four primary interaction estimates after adjustment for HbA1c, insulin, or adiponectin individually or jointly (Table S7).

### 3.6 APOE ε4 allele-dose and genotype sensitivity analyses

APOE ε4 allele-dose and categorical-genotype analyses were examined for the five targeted ventricular and atrial cardiac–brain associations (Table 3; Figure S6). Among participants with ε3/ε3, ε3/ε4, or ε4/ε4 genotypes, all five cardiac phenotype × ε4 allele-dose interactions met q<0.05 after FDR correction.

**Table 3.**
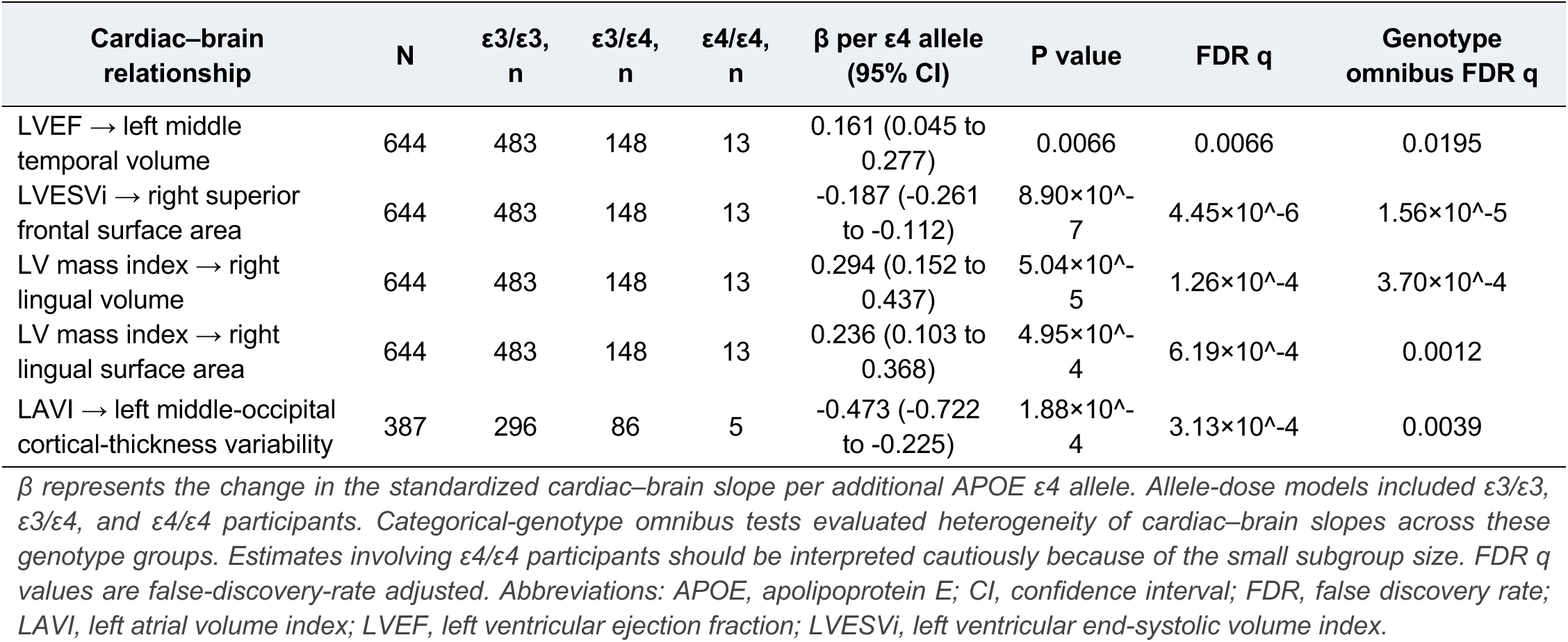
APOE ε4 allele-dose and genotype sensitivity analyses of cardiac–brain associations. Allele-dose models quantify the change in the standardized cardiac–brain slope per additional APOE ε4 allele.

| Cardiac–brain relationship | N | $\epsilon 3/\epsilon 3$, n | $\epsilon 3/\epsilon 4$, n | $\epsilon 4/\epsilon 4$, n | $\beta$ per $\epsilon 4$ allele (95% CI) | P value | FDR q | Genotype omnibus FDR q |
| --- | --- | --- | --- | --- | --- | --- | --- | --- |
| LVEF → left middle temporal volume | 644 | 483 | 148 | 13 | 0.161 (0.045 to 0.277) | 0.0066 | 0.0066 | 0.0195 |
| LVESVi → right superior frontal surface area | 644 | 483 | 148 | 13 | -0.187 (-0.261 to -0.112) | $8.90 \times 10^{-7}$ | $4.45 \times 10^{-6}$ | $1.56 \times 10^{-5}$ |
| LV mass index → right lingual volume | 644 | 483 | 148 | 13 | 0.294 (0.152 to 0.437) | $5.04 \times 10^{-5}$ | $1.26 \times 10^{-4}$ | $3.70 \times 10^{-4}$ |
| LV mass index → right lingual surface area | 644 | 483 | 148 | 13 | 0.236 (0.103 to 0.368) | $4.95 \times 10^{-4}$ | $6.19 \times 10^{-4}$ | 0.0012 |
| LAVI → left middle-occipital cortical-thickness variability | 387 | 296 | 86 | 5 | -0.473 (-0.722 to -0.225) | $1.88 \times 10^{-4}$ | $3.13 \times 10^{-4}$ | 0.0039 |

$\beta$ represents the change in the standardized cardiac–brain slope per additional APOE $\epsilon 4$ allele. Allele-dose models included $\epsilon 3/\epsilon 3$ , $\epsilon 3/\epsilon 4$ , and $\epsilon 4/\epsilon 4$ participants. Categorical-genotype omnibus tests evaluated heterogeneity of cardiac–brain slopes across these genotype groups. Estimates involving $\epsilon 4/\epsilon 4$ participants should be interpreted cautiously because of the small subgroup size. FDR q values are false-discovery-rate adjusted. Abbreviations: APOE, apolipoprotein E; CI, confidence interval; FDR, false discovery rate; LAVI, left atrial volume index; LVEF, left ventricular ejection fraction; LVESVi, left ventricular end-systolic volume index.

For the four ventricular associations, the change in cardiac–brain slope per additional ε4 allele was 0.161 for LVEF–left middle temporal volume (95% CI, 0.045 to 0.277; *P*=0.0066; q=0.0066), −0.187 for LVESVi–right superior frontal surface area (95% CI, −0.261 to −0.112; *P*=8.90×10⁻⁷; q=4.45×10⁻⁶), 0.294 for LV mass index–right lingual volume (95% CI, 0.152 to 0.437; *P*=5.04×10⁻⁵; q=1.26×10⁻⁴), and 0.236 for LV mass index–right lingual surface area (95% CI, 0.103 to 0.368; *P*=4.95×10⁻⁴; q=6.19×10⁻⁴; N=644 for each model). The LAVI–left middle-occipital cortical-thickness-variability association also varied with ε4 allele count (β=−0.473 per additional ε4 allele, 95% CI, −0.722 to −0.225; *P*=1.88×10⁻⁴; q=3.13×10⁻⁴; N=387).

Categorical-genotype interaction tests were also FDR-significant for all five associations (Table 3). Genotype-specific estimates were directionally concordant with the carrier/noncarrier models; for the LAVI association, the allele-dose trend rested on the ε3/ε3 versus ε3/ε4 contrast, and the imprecise ε4/ε4 estimate did not follow a graded pattern (Figure 5C). Estimates for ε4/ε4 participants were interpreted cautiously because of the small subgroup size (n=13 in the ventricular models and n=5 in the atrial model). Because these analyses used the participants in whom the associations were identified, they are directionally consistent sensitivity analyses rather than independent confirmation.

**Figure 5.**
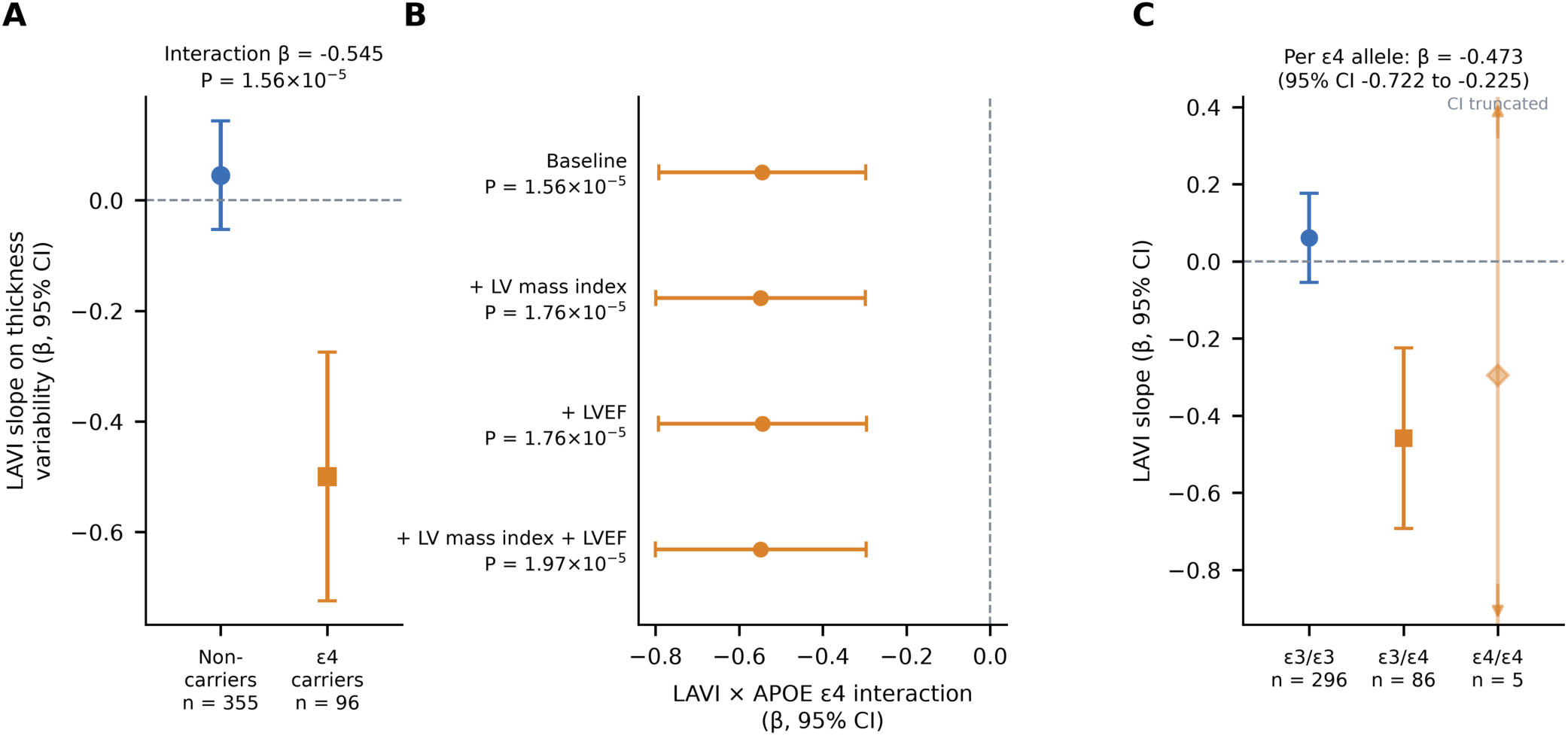
APOE ε4 modifies the association between left atrial volume and left middle-occipital cortical-thickness variability. (A) APOE ε4-stratified standardized slopes of maximum left atrial volume index (LAVI) on left middle-occipital cortical-thickness variability (noncarriers, n=355; carriers, n=96). The LAVI × APOE ε4 interaction was β=−0.545 (95% CI, −0.792 to −0.298; P=1.56×10⁻⁵; whole-brain q=0.017 across 1,086 regional outcomes; whole-screen permutation P=0.041). (B) The interaction after additional adjustment for LV mass index, LVEF, or both, with nominal P values. (C) Genotype-specific LAVI slopes in ε3/ε3 (n=296), ε3/ε4 (n=86), and ε4/ε4 (n=5) participants, with the per-allele interaction (β=−0.473; 95% CI, −0.722 to −0.225); the ε4/ε4 interval is truncated at the axis limits and the estimate is imprecise. Points are coefficients and lines 95% CIs.

### 3.7 APOE ε4 modifies left atrial–brain structural associations

Maximum left atrial volume index (LAVI) showed an APOE ε4-dependent association with left middle-occipital cortical-thickness variability in the primary atrial analysis (N=451; interaction β=−0.545, 95% CI, −0.792 to −0.298; *P*=1.56×10⁻⁵; whole-brain q=0.017 across 1,086 outcomes; whole-screen permutation P=0.041; Figure 5A; Table 4). The estimated standardized LAVI–brain slope was near zero among APOE ε4 noncarriers (β=0.045) and negative among carriers (β=−0.500). No left atrial emptying-fraction × APOE ε4 interaction survived FDR correction.

**Table 4.** LAVI × APOE ε4 interaction after adjustment for ventricular structure and function. Outcome: left middle-occipital cortical-thickness variability; exposure: maximum left atrial volume index (LAVI).

| Model | N | APOE $\epsilon 4$<br>noncarriers,<br>n | APOE<br>$\epsilon 4$<br>carriers,<br>n | $\beta$<br>noncarrier | $\beta$<br>interaction | 95% CI | Interaction<br>P value | $\beta$<br>carrier | R<sup>2</sup> |
| --- | --- | --- | --- | --- | --- | --- | --- | --- | --- |
| Baseline | 451 | 355 | 96 | 0.045 | -0.545 | -0.792 to -0.298 | $1.56 \times 10^{-5}$ | -0.500 | 0.072 |
| + LV mass index | 451 | 355 | 96 | 0.054 | -0.549 | -0.800 to -0.298 | $1.76 \times 10^{-5}$ | -0.495 | 0.073 |
| + LVEF | 451 | 355 | 96 | 0.041 | -0.545 | -0.793 to -0.296 | $1.76 \times 10^{-5}$ | -0.503 | 0.073 |
| + LV mass index + LVEF | 451 | 355 | 96 | 0.050 | -0.549 | -0.801 to -0.297 | $1.97 \times 10^{-5}$ | -0.499 | 0.074 |
| Baseline, all available participants | 476 | 373 | 103 | 0.039 | -0.532 | -0.766 to -0.297 | $8.94 \times 10^{-6}$ | -0.493 | 0.067 |

$\beta$ noncarrier is the standardized LVI–brain slope among APOE $\epsilon 4$ noncarriers. $\beta$ interaction is the difference in slope for APOE $\epsilon 4$ carriers; $\beta$ carrier = $\beta$ noncarrier + $\beta$ interaction. Nested models evaluate attenuation of the LVI × APOE $\epsilon 4$ interaction after additional adjustment for LV mass index, LVEF, or both. Interaction $P$ values are nominal targeted robustness tests. The primary LVI × APOE $\epsilon 4$ association met the FDR criterion in the atrial regional analysis. Abbreviations: APOE, apolipoprotein E; CI, confidence interval; FDR, false discovery rate; LVI, left atrial volume index; LVEF, left ventricular ejection fraction; LV, left ventricular. The last row repeats the baseline model in all 476 participants available for the atrial models; the corresponding adjusted models are in Supplementary Data S2.

The LAVI × APOE ε4 interaction was essentially unchanged after additional adjustment for LV mass index (β=−0.549, 95% CI, −0.800 to −0.298; *P*=1.76×10⁻⁵), LVEF (β=−0.545, 95% CI, −0.793 to −0.296; *P*=1.76×10⁻⁵), or both measures simultaneously (β=−0.549, 95% CI, −0.801 to −0.297; *P*=1.97×10⁻⁵; Figure 5B; Table 4). In a separate analysis using all 476 participants available for the corresponding models, the baseline interaction was similar (β=−0.532; *P*=8.94×10⁻⁶; Table 4). Thus, the LAVI × APOE ε4 interaction showed little attenuation after adjustment for the measured ventricular structural and functional characteristics. It was also preserved under pedigree-family cluster–robust inference (Section 3.8). The interaction was robust to winsorization of the outcome at the 1st and 99th percentiles (β=−0.563; P=2.1×10⁻⁵) and remained significant, although attenuated by 36%, after exclusion of 22 influential observations (11 of 96 carriers and 11 of 355 noncarriers; Cook’s distance >4/N; β=−0.350; P=6.4×10⁻⁴), suggesting that its magnitude may be overestimated.

### 3.8 Sensitivity of primary regional interactions to plasma biomarkers, atrial fibrillation/flutter, and pedigree-family clustering

Adjustment for circulating Aβ42/40, p-tau181, GFAP, and NfL, individually and in prespecified combinations, produced minimal changes in the four primary ventricular cardiac × APOE ε4 interaction estimates (Figure S2; Data S2). Each biomarker-adjusted model was compared with a baseline model fitted to the same participants, separating the effect of adjustment from differences in biomarker availability. Across 28 matched complete-case models (N=678–746), all adjusted estimates remained directionally concordant with their matched-sample baselines, with absolute changes in interaction magnitude of at most 3.2%. These models were interpreted as sensitivity analyses rather than tests of mediation. In a separate analysis of the regional cohort, none of the 16 cardiac phenotype × APOE ε4 interactions with plasma biomarker levels survived FDR correction (minimum q=0.895; Data S2).

The four primary ventricular interactions were similarly preserved after accounting for atrial fibrillation/flutter (AF/flutter) history (Figure S3; Data S2). Adjustment for AF/flutter changed the absolute interaction coefficient by ≤6% for each association, whereas exclusion of participants with AF/flutter changed the coefficients by <8%. For example, the LVEF × APOE ε4 interaction with left middle temporal volume was β=0.234 after adjustment for AF/flutter and β=0.249 after exclusion of participants with AF/flutter, compared with β=0.233 in the primary model. The LVESVi and LV mass index interaction estimates likewise retained their direction and were similar in magnitude to the primary estimates.

Pedigree-family cluster–robust inference provided an additional sensitivity analysis accounting for within-family dependence (Table S7). Models used the fully adjusted covariates with standard errors clustered by pedigree family. All four primary ventricular interactions retained the same direction and remained nominally significant (N=724; 377 pedigrees): LVEF × APOE ε4 with left middle temporal volume (β=0.212; *P*=0.00225), LVESVi × APOE ε4 with right superior frontal surface area (β=−0.134; *P*=0.00368), LV mass index × APOE ε4 with right lingual volume (β=0.319; *P*=1.79×10⁻⁵), and LV mass index × APOE ε4 with right lingual surface area (β=0.210; *P*=0.00119). The LAVI × APOE ε4 association with left middle-occipital cortical-thickness variability was also preserved (β=−0.582; *P*=1.06×10⁻⁷; N=433; 271 pedigrees). In the same samples, HC3 and pedigree-family cluster–robust standard errors were similar (cluster/HC3 ratio, 0.88–0.96) and the coefficients were identical, so differences from the primary coefficients reflect the smaller pedigree-complete samples rather than the variance estimator.

### 3.9 Exploratory metabolic and myocardial-deformation analyses

Exploratory metabolic analyses identified limited evidence of additional APOE ε4-dependent associations (Tables S8–S9). In a focused analysis of HbA1c and LVEDVi, HbA1c showed an APOE ε4-dependent association with LVEDVi (Figure S4; Table S8). In the fully adjusted model, the HbA1c × APOE ε4 interaction was β=−2.89 mL/m² (95% CI, −5.21 to −0.56; *P*=0.0148), with a negative estimated HbA1c slope among APOE ε4 carriers but not among noncarriers. In the broader metabolic analysis, this was the only association within the LV-chamber HbA1c testing family that survived FDR correction. No HbA1c × APOE ε4 or diabetes × APOE ε4 interaction with a brain phenotype survived FDR correction (Table S9).

Exploratory whole-brain analyses of left- and right-ventricular longitudinal, circumferential, and radial strain identified no regional cardiac main effects or cardiac phenotype × APOE ε4 interactions that survived FDR correction (Data S2). Thus, the regional APOE ε4-dependent associations observed for ventricular chamber measures and systolic function were not similarly detected across the myocardial-deformation measures examined at the specified FDR threshold. These null findings do not exclude smaller associations that the study may have been underpowered to detect.

### 3.10 Molecular analyses identify regional enrichment but no convergent pathway-level mechanism

We examined molecular correlates of the four ventricular cardiac × APOE ε4 interactions using participant-level whole-genome sequencing and postmortem reference expression data from the Allen Human Brain Atlas (AHBA). These analyses addressed two distinct questions: whether specific biological processes were enriched within the implicated cortical regions and whether pathway-expression patterns across the cortex corresponded spatially to the cardiac × APOE ε4 interaction maps.

In regional AHBA analyses, genes characteristic of the right superior frontal region implicated by the LVESVi × APOE ε4 surface-area interaction were enriched for the cholesterol biosynthetic process (enrichment ratio=5.60; *P*=1.66×10⁻⁴; FDR q=0.0342; Table S10; Figure S5). Additional FDR-significant regional processes included calcium-ion-regulated exocytosis and regulation of the secretory pathway in right superior frontal cortex and nervous system development in left middle temporal cortex (Table S10). These findings characterize reference molecular profiles of the implicated cortical regions and do not establish that these processes mediate the observed cardiac–brain associations.

We next tested whether inherited variation in seven biological pathway groups modified the four principal cardiac–brain associations among participants with TOPMed whole-genome sequencing data. None of the 28 pathway-level omnibus tests survived FDR correction; the smallest nominal *P* value was observed for the insulin/glucose pathway in the LV mass index–right lingual volume analysis (*P*=0.084; q=0.896; Table S11).

Similarly, targeted AHBA spatial analyses provided no evidence that pathway-expression patterns corresponded to the cortical cardiac × APOE ε4 interaction maps. None of the 28 pathway–map comparisons survived correction using 5,000 Moran spatial-null permutations (minimum spatial *P*=0.319; minimum q=0.914; Data S2). Broader exploratory analyses were also null after global correction, including Gene Ontology Biological Process and Reactome analyses across 17,668 tests (minimum q=0.604; Data S2), MSigDB Hallmark analyses (minimum q=0.963; Data S2), and brain cell-class and more specific cell-signature analyses (Data S2).

Thus, regional reference-expression analyses identified biological-process enrichment within selected implicated cortical regions, including cholesterol biosynthesis in right superior frontal cortex, but the participant-level genetic and cortical spatial analyses did not provide convergent evidence for a pathway or cell-type program underlying the cardiac × APOE ε4 interaction maps. Regional enrichment and spatial correspondence represent distinct analyses and should not be interpreted as equivalent evidence of mechanism.

## 4 Discussion

In the Framingham Heart Study Offspring cohort, APOE ε4 modified the association between antecedent left atrial (LA) remodeling and the rate of subsequent lateral ventricular expansion, the only one of 16 longitudinal heart–brain tests to survive correction for multiple comparisons. Cross-sectionally, maximum left atrial volume index (LAVI) showed an APOE ε4-dependent association with left middle-occipital cortical-thickness variability that survived whole-brain correction and was independent of ventricular measures. Regional associations of left ventricular phenotypes with cortical morphology met within-family FDR criteria but were consistent with chance overall. These findings suggest that APOE ε4 modifies selected, mainly atrial, cardiac–brain relationships rather than producing a uniform association across phenotypes or regions.

These findings extend Framingham observations relating cardiac index and LVEF to brain structure and cognition (Jefferson et al. 2010; Jefferson et al. 2011) and evidence that cardiac–brain associations are phenotype- and region-specific (Zhao et al. 2023). (Moore et al. 2026) reported more adverse associations of lower cardiac output with brain change among APOE ε4 carriers, and the exploratory LVEF and LVESVi interactions here agreed in direction. The longitudinal atrial finding did not: faster LA enlargement was associated with faster ventricular expansion in noncarriers, whereas this association was absent in carriers. Conversely, left atrial volume was associated with occipital cortical-thickness variability only in carriers. The studies are not directly comparable, because Moore et al. related baseline cardiac output to inferior lateral ventricular change in a different cohort, but together these results do not support a model in which APOE ε4 uniformly amplifies adverse cardiac–brain associations. Rather, they raise the hypothesis that atrial-related brain vulnerability in carriers is regional rather than global: the association with global ventricular expansion was absent, whereas a focal cortical association emerged. Because the two analyses used different exposures and designs, this pattern requires direct testing.

Several explanations for the absent longitudinal association in carriers deserve consideration. First, ventricular expansion is faster in ε4 carriers (Roussotte et al. 2014) and may be dominated by APOE ε4-related neurodegenerative processes, diluting a hemodynamic contribution of atrial remodeling. Second, selective attrition: carriers with both rapid atrial remodeling and rapid brain decline may have been less likely to undergo brain MRI. Third, APOE ε4 is associated with cardiac physiology: carriers have shown higher myocardial contraction fraction and more favorable strain rates (Topriceanu et al. 2024), so equal values of a cardiac measure may reflect different hemodynamic states across genotypes. Experimental studies suggest that APOE isoform influences cardiomyocyte contractility and calcium handling (Wu et al. 2023), and APOE ε4 has been associated with impaired endothelium-dependent cerebrovascular responses and neurovascular coupling (Koizumi et al. 2018) and with reduced cerebrovascular reactivity (Suri et al. 2015), which could change how a given cardiac state translates into cerebral perfusion. These processes were not measured here and could act in either direction. The longitudinal LA finding provides the clearest temporal ordering: LA remodeling was measured across serial examinations that preceded repeated brain MRI. The point estimate was similar with age- and sex-dependent trajectories, participant-specific random MRI-time slopes, adjustment for baseline LA dimension, and an Examination 4–6 definition of remodeling; with random slopes it remained the only FDR-significant longitudinal interaction. The generalized estimating equation estimate was similar in magnitude but less precise (P=0.040). The interaction was smaller in the ICV-complete subset before ICV terms were added, indicating that sample restriction accounted for much of the attenuation, and inclusion of ICV × MRI time yielded a confidence interval that narrowly included the null. Native-unit predictions showed ventricular expansion in both groups, so the interaction reflects a genotype-dependent difference in the rate of expansion, not contraction among carriers.

Among the exploratory regional associations, the LVESVi–right superior frontal interaction was preserved when CMR and brain MRI were obtained within three or two years, whereas the LV mass–right lingual associations attenuated toward the null. The implicated cortical regions do not form a single Alzheimer disease-specific or vascular-territory pattern. Superior frontal cortex belongs to the cortical signature of Alzheimer disease, in which thinning appears later than in medial temporal and inferior parietal cortex (Dickerson et al. 2009), whereas middle temporal and lingual cortex lie outside it; this correspondence is approximate. APOE ε4-related blood–brain barrier dysfunction, described mainly in the hippocampus and medial temporal lobe (Montagne et al. 2020), provides one plausible context for genotype-dependent cerebral associations, but perfusion, barrier permeability, and microinfarction were not assessed, so the anatomical findings generate neurovascular hypotheses only.

The atrial analyses address distinct exposures and outcomes. CMR-derived LAVI was related to left middle-occipital cortical-thickness variability; the interaction showed little attenuation after adjustment for LVEF and LV mass and, unlike the regional ventricular findings, survived correction across all 1,086 outcomes of the atrial screen by both FDR and whole-screen permutation (P=0.041). Cortical-thickness variability reflects within-region heterogeneity of thickness, which can be altered by focal thinning or microstructural change, and the occipital cortex lies in the posterior circulation, a territory relevant to atrial-related embolic and hemodynamic injury; neither mechanism was assessed here. The association was attenuated after exclusion of influential observations, and thickness variability is sensitive to segmentation and partial-volume effects, so confirmation with other morphometric measures is needed. LA remodeling may reflect cumulative filling pressure, vascular load, or atrial myopathy, and atrial abnormalities have been linked to brain injury and dementia (Austin et al. 2022) (Wang et al. 2022). Clinically recognized atrial fibrillation or flutter did not explain the regional ventricular interactions, although occult arrhythmia and embolic injury remain possible.

APOE ε4 allele-dose and categorical-genotype analyses were directionally concordant with the regional associations, but, because they used the same participants, they cannot offset the permutation result. For the longitudinal atrial association, genotype-specific slopes differed overall but did not follow ε4 allele count. Neither analysis constitutes independent replication, because both used the same cohort and the ε4/ε4 subgroups were small. In matched complete cases, adjustment for plasma Aβ42/40, p-tau181, GFAP, and NfL changed the principal ventricular interaction estimates by at most 3.2%, and the ventricular and atrial interactions remained nominally significant with pedigree-family cluster–robust standard errors. Neither cardiac phenotypes nor antecedent cardiac trajectories showed FDR-corrected APOE ε4-dependent associations with plasma biomarkers, and no interaction survived correction across the later cognitive-test analyses. The one FDR-significant cognitive result, an LV mass trajectory × time × APOE ε4 interaction for repeated MMSE, involved a different cardiac exposure from the principal atrial finding and a coarse global measure, and it requires replication. The structural findings therefore should not be interpreted as evidence of corresponding biomarker or cognitive change during the available follow-up.

Among exploratory metabolic analyses, current diabetes status showed no FDR-significant APOE ε4-dependent associations, whereas higher HbA1c was associated with smaller LV end-diastolic volume index among ε4 carriers but not noncarriers (interaction P=0.015; Table S8). This signal emerged from a screen and involved a cardiac rather than a brain outcome, and suggests that glycemic exposure may contribute to genotype-dependent cardiac remodeling. Molecular analyses were likewise hypothesis-generating: Allen Human Brain Atlas data identified cholesterol-biosynthesis enrichment in the implicated right superior frontal region, consistent with the role of APOE in lipid transport, but participant-level genetic and cortical spatial analyses did not provide convergent pathway evidence.

To our knowledge, this is the first study to test whether APOE ε4 modifies associations of left atrial, as distinct from ventricular, phenotypes with brain structure, and to do so with cardiac remodeling measured before more than a decade of repeated brain MRI.

Several limitations should be considered. The regional ventricular findings met FDR criteria only within testing families that were finalized after initial analyses, and their number was consistent with chance; they require replication with prespecified models. Adjustment for antihypertensive treatment at Examination 4 did not change the longitudinal interaction, but treatment class and changes over time were not examined. The observational design precludes causal inference, and CMR and brain MRI were obtained at separate visits in the regional analyses. Longitudinal analyses required survival and continued participation; inclusion rates were similar across APOE ε4 status and LA-remodeling tertiles, the interaction did not predict repeated-MRI inclusion, and selection weighting gave a similar estimate, but selection on unmeasured factors cannot be excluded. Atrial remodeling was estimated from three M-mode LA dimensions, a limited measure subject to measurement error. Complete-case samples varied across analyses, ε4/ε4 subgroups were small (5 to 23 participants), and incompletely measured vascular exposures may have contributed. Cortical morphometry and ventricular volume are not specific to Alzheimer disease, and transcriptomic and germline analyses cannot establish mediation. Finally, the predominantly non-Hispanic White Framingham Offspring cohort limits generalizability.

Despite these limitations, if replicated, these findings would identify left atrial remodeling, measurable on routine echocardiography, as a marker of brain aging whose relevance depends on APOE genotype, and they argue for genotype-stratified analyses in heart–brain studies, in which pooled estimates may obscure associations that differ in form between carriers and noncarriers. They also motivate longitudinal tests of whether atrial remodeling and glycemic exposure act on the brain through cerebral perfusion, and of whether APOE genotype modifies the incidence of heart failure with preserved ejection fraction.

## 5 Conclusion

In the Framingham Offspring cohort, APOE ε4 modified the association between antecedent left atrial remodeling and subsequent lateral ventricular expansion, and left atrial volume showed an APOE ε4-dependent association with occipital cortical-thickness variability, whereas regional ventricular associations were exploratory. APOE ε4 may alter the form of heart–brain relationships rather than uniformly amplifying adverse associations. Confirmed in independent cohorts, these findings would position left atrial remodeling, which is routinely measured and shaped by modifiable factors such as blood pressure, rhythm, and glycemia, as an accessible, genotype-informed window on brain aging.

## Acknowledgments

The authors thank the participants and investigators of the Framingham Heart Study for their longstanding contributions to research. The Framingham Heart Study is conducted and supported by the National Heart, Lung, and Blood Institute (NHLBI) in collaboration with Boston University and is supported by NHLBI contracts 75N92025D00012 and 75N92019D00031, with additional support from N01-HC-25195, HHSN268201500001I, R01HL107385, and U19AG023122.

Data used in this study were obtained through the NIH database of Genotypes and Phenotypes (dbGaP), including the Framingham Cohort (phs000007.v35.p16), NHLBI TOPMed: Genomic Activities such as Whole Genome Sequencing and Related Phenotypes in the Framingham Heart Study (phs000974.v6.p5), and the Framingham Heart Study–Cohort Imaging study (phs003593.v2.p1). In accordance with the study-specific acknowledgment guidance associated with the Framingham imaging resource, we additionally acknowledge the Framingham Cohort accession phs000007.v33.p14.

This manuscript was not prepared in collaboration with investigators of the Framingham Heart Study and does not necessarily reflect the opinions or conclusions of the Framingham Heart Study, Boston University, NHLBI, or the National Institutes of Health.

Claude (Anthropic) and ChatGPT (OpenAI) were used to assist with analysis code, figure generation, reproducibility checks, and language editing. The authors reviewed and verified all analyses, results, and text and take full responsibility for the content.

## Data Availability Statement

Framingham Heart Study data used in this study are available to qualified investigators through the NIH database of Genotypes and Phenotypes (dbGaP) under controlled access: the Framingham Cohort (phs000007.v35.p16), NHLBI TOPMed whole-genome sequencing in the Framingham Heart Study (phs000974.v6.p5), and the Framingham Heart Study–Cohort Imaging study (phs003593.v2.p1). Under the terms of the data-use agreement, participant-level data cannot be shared by the authors. Allen Human Brain Atlas gene-expression data are publicly available from the Allen Institute for Brain Science (https://human.brain-map.org). Complete summary-level results, including all regional tests and the values underlying the supplementary figures, are provided in Supplementary Data S1 and S2. Analysis code, including scripts that regenerate all figures and tables and verify them against the reported values, is available at https://github.com/AD-Decode/atrial_brain.

