## SupplementaryMaterial for "APOE ε4 Modifies the Association of Left Atrial Remodeling With Brain Aging"

#### Contents

Supplementary Methods

Table S1. Antecedent left atrial remodeling, APOE ε4, and subsequent lateral ventricular change: principal interaction, carrier-stratified slopes, and APOE genotype

Table S2. Complete longitudinal testing family: antecedent cardiac trajectory × MRI time × APOE ε4 interactions, random-intercept and random-slope models

Table S3. Sensitivity of the LA remodeling × MRI time × APOE ε4 association to longitudinal model specification, exposure definition, head size, and early vascular covariates

Table S4. Availability of subsequent brain MRI, selection into the repeated-MRI cohort, and inverse-probability-of-selection weighting

Table S5. Antecedent cardiac trajectory × APOE ε4 interactions with later plasma biomarkers and later cognition

Table S6. Regional cardiac–brain associations meeting the FDR criterion for the cardiac main effect or the cardiac × APOE ε4 interaction

Table S7. Sensitivity of the four regional ventricular interactions to metabolic markers, and pedigree-family cluster–robust inference

Table S8. APOE ε4 modification of the association between HbA1c and LV end-diastolic volume index

Table S9. Exploratory APOE ε4 × metabolic-modifier analyses across brain and cardiac phenotype families

Table S10. Biological processes enriched in implicated cortical regions in the Allen Human Brain Atlas reference data

Table S11. Summary of molecular analyses for the four regional ventricular associations

Supplementary Figures S1–S6

Supplementary Data S1 (separate Excel workbook): complete regional M1, M2, and M3 results.

Supplementary Data S2 (separate Excel workbook): numerical values underlying Figures S1–S6, and detailed results moved from the print supplement (stratified slopes, LAVI models in all available participants, plasma-biomarker and AF/flutter sensitivity models, cross-sectional biomarker associations, three-dimensional strain analyses, and whole-cortex spatial-correspondence tests).

### Supplementary Methods

#### Cohort construction and examination timing

The regional imaging-overlap cohort paired one CMR assessment and one brain MRI assessment per participant. The initial overlap numbered 785; 769 participants had classified APOE genotype. Fully adjusted regional models used model-specific complete cases (765 for the four principal ventricular associations). CMR and brain MRI were separated by a mean absolute interval of 2.96 years (SD, 1.26; range, 0.008–7.72). The absolute elapsed time entered the model as a covariate; targeted sensitivity subsets restricted this interval to at most three and at most two years. The longitudinal arm used echocardiography from Offspring Examinations 4–6, generally approximately 3–4 years apart, followed by repeated MRI. The exact Exam 4–5 and 5–6 gaps were not assumed to be equal in the slope calculation. Each outcome-specific primary analysis required three nonmissing cardiac measurements, a valid cardiac slope, APOE classification, age at Examination 6, sex, a nonnegative interval from the final cardiac assessment to the first eligible MRI, and at least two eligible MRI outcomes. The two-year-gap sensitivity retained only participants whose first eligible MRI occurred at least two years after that assessment. In the principal longitudinal lateral-ventricle analysis, the median last-echo-to-first-MRI interval was 3.47 years (IQR, 2.83–4.28), with median MRI follow-up of 12.45 years (IQR, 6.84–13.94) and a median of three MRI observations per participant (IQR, 2–3). Sample sizes were recalculated for every cardiac-outcome combination.

#### Cardiac measures and trajectory estimation

The Offspring CMR examinations were acquired during 2002–2006 on a 1.5-T system using cine steady-state free-precession imaging; published Framingham protocols describe contiguous 10-mm short-axis LV images and contour-based estimates of LV volumes, mass, and ejection fraction. CMR-derived LV end-diastolic and end-systolic volumes and LV mass were indexed to body surface area, calculated as $\sqrt{[\text{height (cm)} \times \text{weight (kg)} / 3600]}$; LVEF was retained as a percentage. Indexed LAVI, atrial functional measures, and three-dimensional LV and right ventricular strain were considered in supporting analyses. Serial echocardiographic traits were left atrial dimension, LV end-diastolic dimension, derived LV mass, and fractional shortening. Harmonized examination dates were used for the exposure window. The participant-level slope for each cardiac measure was the coefficient of a straight-line least-squares fit of the observed measurement against years elapsed since the first dated examination. The source code uses np.polyfit(x, y, 1)[0]; the slope is measured in the native cardiac unit per year. Although trajectory construction permits a slope with two dated observations, the principal longitudinal models explicitly require all three Exam 4–6 measurements. Cardiac slopes were centered and divided by their participant-level standard deviation separately within each eligible model sample.

#### Regional brain measures and model scaling

Framingham T1-weighted brain MRI was acquired using a 1.5-T Siemens scanner; FreeSurfer-derived morphometry was obtained from dbGaP dataset pht004364. Structural MRI supplied hemisphere-specific Desikan–Killiany and Destrieux measures of volume, surface area, and thickness. Regional volumes were divided by intracranial volume (ICV) and then standardized. Alternative volume models used unscaled regional volume with ICV as a separate covariate. Surface area was standardized without ICV division, with standardized total cortical surface area entered as a covariate. Thickness was not divided by ICV. In addition to mean cortical thickness, regional cortical thickness standard deviation (TKSD, mm), as provided in the Framingham FreeSurfer-derived morphometric dataset, was analyzed as a measure of within-region cortical thickness variability. Continuous cardiac exposures, outcomes, age, CMR–MRI interval, BMI, and systolic pressure were standardized within the complete-case regional model. Sex, current smoking, diabetes history, and APOE ε4 status retained their categorical coding.

#### Regional discovery and correction families

The primary APOE ε4 indicator distinguished carriers from noncarriers. Follow-up categorical-genotype analyses used ε3/ε3 as the reference genotype; estimates for ε4/ε4 participants were interpreted cautiously because this subgroup was small. Eligible regions were assigned to eight anatomical families: seven cortical families (medial temporal/Alzheimer disease-relevant, cingulate, insula, frontal, parietal/precuneus, other temporal, and occipital), with hippocampal and amygdalar volumes assigned to the medial temporal family, and one subcortical-nuclei family (caudate, putamen, pallidum, thalamus, and nucleus accumbens; volume only). The family structure was finalized after initial analyses. Assignment was by label matching: 28 Desikan–Killiany and 35 Destrieux regions per hemisphere matched a family and were analyzed for volume, surface area, and thickness, together with seven bilateral subcortical volumes (392 outcomes per cardiac phenotype). Six Desikan–Killiany regions per hemisphere (banks of the superior temporal sulcus, pars opercularis, pars orbitalis, pars triangularis, paracentral, and supramarginal) and 39 Destrieux regions per hemisphere matched no family and were not tested; the excluded labels are listed in the code repository. The atrial screen used a different, suffix-based rule: all labels ending in volume, surface-area, thickness, thickness-variability, or mean-curvature suffixes, comprising 68 Desikan–Killiany and 148 Destrieux labels × 5 metrics (1,080 outcomes), four hemispheric white-surface-area summaries, and the left and right accumbens volumes, whose FreeSurfer label (Accumbens-area) ends in "area" (1,086 outcomes in total). These six variables were retained so that the analysis and its permutation test remained as originally run. Desikan–Killiany and Destrieux parcels assigned to the same family entered the same testing group. The fully adjusted regional model (M3) included the standardized cardiac measure, APOE ε4 status, their interaction, age at brain MRI, sex, absolute CMR–MRI interval, body mass index, systolic blood pressure, smoking, and diabetes. Regional volumes were divided by intracranial volume before standardization; surface-area models additionally adjusted for total cortical surface area. Continuous variables were standardized within model-specific complete cases, and ordinary-least-squares models used HC3 robust standard errors. Benjamini–Hochberg correction was applied to interaction P values separately within each brain metric × anatomical family × cardiac phenotype testing group. Reported q values therefore refer to these groups, rather than to a single correction across the entire regional screen. As an exploratory sensitivity analysis, Benjamini–Hochberg correction was also examined across the full regional interaction screen; no interaction survived this global correction (minimum global q=0.21). The complete screen was also evaluated with a Freedman–Lane permutation test (5,000 permutations; seed 20261006). Residuals from the fully adjusted M3 model without the cardiac × APOE ε4 term were permuted with a single permutation applied jointly to all 1,960 outcomes, preserving the APOE ε4 main effect, covariate structure, and correlation among regions; refitted observed P values reproduced the reported values. The observed number of within-family discoveries (4) did not exceed the null expectation (null median, 6; mean, 11.6; 95th percentile, 40; permutation P=0.70), at least one within-family discovery occurred in 95% of permutations, and the smallest observed interaction P value was consistent with chance (minimum-P permutation P=0.28). In a post hoc focused non-redundant subset of 546 tests (28 eligible Desikan–Killiany cortical regions per hemisphere plus hippocampal, amygdalar, and subcortical nuclear volumes; LVEF, LVESVi, and LV mass index; all three morphometric types), the minimum-P permutation P was 0.11. In an earlier cortical-only subset of 504 tests (28 eligible Desikan–Killiany cortical regions per hemisphere; LVEF, LVESVi, and LV mass index; volume, surface area, and thickness), the overall minimum-P permutation P was 0.10. Metric-specific minimum-P permutation P values were 0.043 for volume, 0.045 for surface area, and 0.801 for thickness (168 tests each); after Benjamini–Hochberg correction across the three metrics, q=0.068, 0.068, and 0.801, respectively. These focused analyses were specified after the complete-screen result. The LAVI association, analyzed in a separate whole-brain atrial screen of 1,086 regional outcomes, had q=0.017; in a 5,000-permutation Freedman–Lane minimum-P test of that screen, preserving outcome-specific complete cases (refitted P values reproduced the originals), the empirical whole-screen P was 0.041. Because this exceeds the Bonferroni bound computed from the parametric P value (0.017), the parametric HC3 P value for this interaction is likely somewhat anti-conservative in the extreme tail, and the permutation P is the preferred familywise estimate. Accordingly, the regional ventricular findings are interpreted as exploratory and hypothesis-generating rather than as globally FDR-significant associations. Regional associations meeting the FDR criterion are listed in Table S6; complete results for all models are provided in Supplementary Data S1.

#### Targeted regional robustness analyses

For the four ventricular associations, targeted analyses compared the original model with subsets having CMR–MRI intervals of at most three and two years, raw-volume/ICV-covariate specifications where applicable, APOE ε4 allele-dose and categorical-genotype models, pedigree-family cluster–robust standard errors, adjustment for atrial fibrillation/flutter history, and exclusion of participants with such history. For the interval-restricted analyses displayed in Figure 4, standardization parameters were estimated from the corresponding full-cohort model-specific complete-case sample and held fixed in the ≤3-year and ≤2-year subsets, placing coefficients from the restricted analyses on the same scale as the corresponding full-cohort estimate. Other supporting analyses assessed alternative systolic and diastolic blood-pressure specifications, antihypertensive treatment, metabolic covariates including glycemic measures, and plasma biomarker adjustment. The LAVI × APOE ε4 association with occipital cortical-thickness variability was tested after additional adjustment for LVMI, LVEF, or both. Some targeted associations weakened under restrictions; follow-up P values are therefore reported with sample sizes and are not interpreted as independent confirmation.

#### Longitudinal brain models and robustness

The longitudinal MRI outcomes were hippocampal volume, total brain volume, lateral ventricular volume, and white matter hyperintensity burden. For each eligible cardiac trajectory–brain outcome sample, MRI time was defined as years since the first eligible post-cardiac MRI. Cardiac trajectories were standardized across eligible participants, and brain outcomes were standardized using the mean and standard deviation across eligible MRI observations; standardization therefore depended on the model-specific analytic sample.

The principal analysis used maximum-likelihood linear mixed-effects models with a participant-specific random intercept and the specification brain_z ~ mri_time_years * cardiac_z * APOE4_carrier + age6_c + C(sex). All constituent lower-order interaction terms were included. Age at Examination 6 was centered. The cardiac trajectory × MRI time coefficient estimated the trajectory association among APOE ε4 noncarriers, whereas the cardiac trajectory × MRI time × APOE ε4 coefficient estimated the carrier-minus-noncarrier difference in that association. The principal model did not include age × MRI time or sex × MRI time interactions or a participant-specific random MRI-time slope.

Benjamini–Hochberg FDR correction was applied separately within the prospective and ≥2-year-gap subsets to the 16 cardiac trajectory × MRI time × APOE ε4 tests. Carrier-stratified slopes, vascular and metabolic adjustment, generalized estimating equations (GEE), and APOE genotype analyses restricted to ε3/ε3, ε3/ε4, and ε4/ε4 participants (ε3/ε3 reference; 2-df Wald test of overall genotype modification) were targeted follow-up analyses. GEE yielded a similar point estimate for the principal prospective interaction but greater uncertainty, and the interaction did not survive its corresponding FDR correction; GEE was therefore interpreted as a robustness analysis rather than an independently multiplicity-significant finding. Principal, carrier-stratified, GEE, and APOE genotype estimates for the LA remodeling–lateral ventricular association are reported in Table S1, the complete 16-test family in Table S2, and model-specification, head-size, and early vascular sensitivity analyses in Table S3.

Targeted sensitivity analyses of the principal LA remodeling–lateral ventricular association evaluated longitudinal model specification and exposure definition. First, age at Examination 6 × MRI time and sex × MRI time interactions were added to allow longitudinal brain trajectories to vary with baseline age and sex. A subsequent model incorporated a participant-specific random MRI-time slope in addition to the random intercept. The expanded model additionally included baseline LA dimension and baseline LA dimension × MRI time while retaining the age × MRI time and sex × MRI time terms and participant-specific random MRI-time slope.

To evaluate sensitivity to estimation of the antecedent remodeling exposure, an alternative LA remodeling measure was calculated directly from Examination 4 and Examination 6 measurements as the change in LA dimension divided by the elapsed time between the dated examinations. Examination 4 LA dimension and its interaction with MRI time were included in this analysis. This specification therefore tested whether the longitudinal finding depended on fitting the principal multi-examination LA trajectory.

Head-size sensitivity analyses were performed in participants with available ICV. Because ICV availability reduced the analytic sample, comparisons were made within the same ICV-complete subset. Matched random-intercept models were fitted without ICV terms, with ICV as an additional covariate, and with both ICV and ICV × MRI time. The latter specification allowed the rate of ventricular-volume change to vary with head size. A corresponding random-slope ICV specification did not converge and was not interpreted.

For native-unit interpretation, the standardized LA remodeling × MRI time × APOE ε4 interaction from the expanded model was back-transformed using the observed standard deviations of the LA remodeling trajectory and total lateral ventricular volume in its analytic sample. LA remodeling was expressed in cm/year and total lateral ventricular volume in mL. Predicted annual ventricular-volume change was evaluated across the observed LA-remodeling range separately for APOE ε4 carriers and noncarriers. These analyses were used to distinguish an interaction in the rate of ventricular expansion from ventricular contraction. Full longitudinal sensitivity estimates are reported in Table S3.

##### Selection into the repeated-MRI cohort

The pre-MRI at-risk cohort comprised participants with a valid three-examination LA remodeling trajectory and classified APOE genotype, defined before conditioning on subsequent MRI participation. Inclusion was defined as at least two eligible lateral ventricular MRI observations after the final cardiac examination. MRI availability was summarized overall and by APOE ε4 status and tertile of LA remodeling (Table S4). Inclusion was modeled by logistic regression on standardized LA remodeling, APOE ε4 status, their interaction, age at Examination 6, sex, and Examination 4 body mass index, systolic blood pressure, diabetes, smoking, and LA dimension. Stabilized inverse-probability-of-selection weights were truncated at the 1st and 99th percentiles, and the LA remodeling × MRI time × APOE ε4 interaction was re-estimated by weighted GEE and compared with an unweighted GEE in the same participants (Table S4).

##### Plasma biomarkers

Plasma amyloid-β42/40 (Aβ42/40), phosphorylated tau 181 (p-tau181), glial fibrillary acidic protein (GFAP), and neurofilament light chain (NfL) entered three distinct analyses. First, the four primary ventricular cardiac × APOE ε4 brain models were refitted with each biomarker individually and with three prespecified combinations (Aβ42/40 + p-tau181; GFAP + NfL; all four). Each adjusted model was compared with the M3 model fitted to the identical complete-case sample, separating adjustment effects from changes in sample membership; FDR was calculated across the four associations within each specification (Data S2). Second, in the regional cohort, cardiac phenotype × APOE ε4 interactions were tested with each biomarker as the outcome, using ordinary least-squares models with HC3 standard errors adjusted for age, sex, body mass index, systolic blood pressure, smoking, and diabetes; p-tau181, GFAP, and NfL were log-transformed, outcomes and cardiac phenotypes were standardized, and FDR was calculated across the 16 tests (Data S2). Third, standardized Examination 4–6 cardiac trajectories were related to later biomarker outcomes, with FDR within each cardiac trajectory and across all 16 tests (Table S5). Assay-specific handling followed the relevant Framingham data dictionaries.

#### Cognition

Repeated later MMSE-based scores, normalized by the maximum available for each administration, were analyzed in participants with at least two eligible post-exposure administrations. For each of the four cardiac trajectories, a longitudinal model estimated the cardiac trajectory × cognitive time × APOE ε4 interaction, with FDR across the four tests (Table S5, Panel B). Four later neuropsychological tests (Logical Memory delayed recall, Trails B, animal fluency, and Digit Span Backward) were analyzed at each participant's first eligible later assessment as exploratory outcomes. Cardiac trajectory × APOE ε4 interactions were estimated for all 16 trajectory × test combinations, with FDR across the 16 tests as the primary correction family and within-trajectory q values reported for reference (Table S5, Panel A).

#### Participant-level WGS and AHBA spatial analyses

The participant-level whole-genome sequencing analysis tested seven biological gene-set groups for cardiac effect modification in each of the four principal ventricular cardiac–brain associations (28 omnibus tests; Table S11). The AHBA analyses used postmortem reference expression rather than expression measured in Framingham participants. The whole-cortex interaction maps were constructed from regional M3 cardiac × APOE ε4 estimates, aligned with the Desikan–Killiany cortical atlas. AHBA expression was processed with abagen using differential-stability probe selection, bidirectional mirroring, interpolation of missing parcels, scaled robust sigmoid sample and gene normalization, within-structure normalization, and donor-level aggregation. The targeted analysis examined seven pathway-expression scores against each of four interaction maps across 56 cortical parcels. Spearman correspondence was evaluated against 5,000 Moran spatial surrogate maps, followed by FDR correction across the 28 targeted comparisons (Data S2). Exploratory spatial analyses assessed GO Biological Process and Reactome terms, MSigDB Hallmark sets, and brain cell-class and cell-signature sets (all in Data S2). Within-map and global FDR values were reported for their respective analysis families. Table S11 synthesizes genetic and spatial analyses; no corrected pathway convergence was established.

#### AHBA regional enrichment and provenance

The reference gene-expression measurements came from the Allen Institute for Brain Science's Allen Human Brain Atlas adult postmortem microarray resource (https://human.brain-map.org/static/download). A region-level DK83 reference atlas derived from these measurements provided the enrichment results displayed in Table S10 and Figure S5. The implicated left middle temporal, right superior frontal, and right lingual regions were selected for display. The regional reference-atlas enrichment and the whole-cortex AHBA spatial-correspondence tests are distinct analyses. The right superior frontal cholesterol biosynthetic process enrichment (GO:0006695; ratio 5.60; q=0.0342) reflects enrichment within that reference region; it does not establish spatial correspondence of cholesterol expression with the cardiac–brain interaction map.

#### Family structure, missing data, and interpretation

Models used analysis-specific complete cases. Pedigree-family cluster–robust standard errors were calculated as targeted regional sensitivity analyses rather than by kinship-matrix or pedigree random-effects modeling. HC3 standard errors from the same pedigree-complete samples were computed for comparison (Table S7). Results were reported with participant and observation counts, coefficient scales, confidence intervals, nominal P values, and their corresponding correction families. Prospective measurement order does not establish causality.

#### Table S1. Antecedent left atrial remodeling, APOE ε4, and subsequent lateral ventricular change: principal interaction, carrier-stratified slopes, and APOE genotype

***Panel A. LA remodeling × MRI time × APOE ε4 interaction***

| Sample | Method | N | MRI observations | Carriers / noncarriers, n | β (95% CI) | P | FDR q |
|---|---|---|---|---|---|---|---|
| Prospective | Mixed model | 1,305 | 3,787 | 303 / 1,002 | −0.00858 (−0.01300 to −0.00415) | $1.45 \times 10^{-4}$ | 0.00231 |
| Prospective | GEE | 1,305 | 3,787 | 303 / 1,002 | −0.00852 (−0.01666 to −0.00039) | 0.0399 | 0.368 |
| ≥2-year gap | Mixed model | 1,260 | 3,646 | 288 / 972 | −0.00813 (−0.01270 to −0.00356) | $4.84 \times 10^{-4}$ | 0.00774 |
| ≥2-year gap | GEE | 1,260 | 3,646 | 288 / 972 | −0.00807 (−0.01660 to 0.00046) | 0.0637 | 0.431 |

***Panel B. APOE ε4-stratified LA remodeling × MRI time slopes***

| Sample | Method | APOE ε4 group | N | Slope β (95% CI) | P |
|---|---|---|---|---|---|
| Prospective | Mixed model | APOE ε4 noncarrier | 1,305 | 0.00357 (0.00147 to 0.00567) | $8.78 \times 10^{-4}$ |
| Prospective | Mixed model | APOE ε4 carrier | 1,305 | −0.00501 (−0.00890 to −0.00112) | 0.0117 |
| Prospective | GEE | APOE ε4 noncarrier | 1,305 | 0.00356 (0.00078 to 0.00633) | 0.0121 |
| Prospective | GEE | APOE ε4 carrier | 1,305 | −0.00497 (−0.01261 to 0.00268) | 0.203 |
| ≥2-year gap | Mixed model | APOE ε4 noncarrier | 1,260 | 0.00314 (0.00100 to 0.00528) | 0.00400 |
| ≥2-year gap | Mixed model | APOE ε4 carrier | 1,260 | −0.00499 (−0.00902 to −0.00095) | 0.0154 |
| ≥2-year gap | GEE | APOE ε4 noncarrier | 1,260 | 0.00312 (0.00036 to 0.00588) | 0.0266 |
| ≥2-year gap | GEE | APOE ε4 carrier | 1,260 | −0.00495 (−0.01301 to 0.00312) | 0.230 |

***Panel C. APOE genotype (participants with ε3/ε3, ε3/ε4, or ε4/ε4; prospective sample)***

| Genotype / test | n | LA remodeling × MRI time β | P |
|---|---|---|---|
| ε3/ε3 (reference) | 823 | +0.00211 | 0.0700 |
| ε3/ε4 | 256 | −0.00552 | 0.00910 |
| ε4/ε4 | 23 | +0.00622 | 0.442 |
| Joint test of genotype modification (2-df Wald) | 1,102 | $\chi^2 = 10.50$ | 0.00525 |

Exposure: participant-specific slope of echocardiographic LA dimension across Offspring Examinations 4–6, standardized within each analytic sample. Outcome: total lateral ventricular volume across subsequent MRI examinations, standardized across eligible observations. Coefficients are in standardized brain-outcome units per year per SD of LA remodeling.

Mixed models used a participant-specific random intercept and adjusted for age at Examination 6 and sex. GEE used an exchangeable working correlation and robust standard errors. FDR q values were calculated across the 16 trajectory × outcome tests within each sample and method.

Panel B: noncarrier slopes are the LA remodeling × MRI time coefficient; carrier slopes add the three-way interaction. Panel C: restricted to ε3/ε3, ε3/ε4, and ε4/ε4 participants, with the same specification as the primary carrier model and ε3/ε3 as the reference; genotype-specific estimates are the LA remodeling × MRI time effects within each genotype. The joint test assesses whether these effects differ across genotypes; the pattern is not monotonic in ε4 allele count. The ε4/ε4 estimate is imprecise (n=23).

Vascular-adjusted estimates are in Table S3. Abbreviations: APOE, apolipoprotein E; CI, confidence interval; FDR, false discovery rate; GEE, generalized estimating equations; LA, left atrial.

#### Table S2. Complete prospective longitudinal testing family: antecedent cardiac trajectory × MRI time × APOE ε4 interactions for four subsequent brain outcomes

| Cardiac trajectory | Brain outcome | N | MRI observations | Carriers / noncarriers, n | Interaction β (95% CI) | P | FDR q |
|---|---|---|---|---|---|---|---|
| LA dimension | Lateral ventricular volume | 1,305 | 3,787 | 303 / 1,002 | −0.00858 (−0.01300 to −0.00415) | $1.45\times10^{-4}$ | 0.00231 |
| Fractional shortening | Lateral ventricular volume | 952 | 2,812 | 235 / 717 | 0.00570 (0.00106 to 0.01034) | 0.0161 | 0.108 |
| Fractional shortening | White matter hyperintensity burden | 630 | 1,395 | 154 / 476 | −0.01727 (−0.03186 to −0.00268) | 0.0203 | 0.108 |
| Fractional shortening | Total brain volume | 952 | 2,812 | 235 / 717 | 0.00252 (−0.00092 to 0.00595) | 0.151 | 0.566 |
| LV mass | White matter hyperintensity burden | 638 | 1,415 | 156 / 482 | 0.01140 (−0.00551 to 0.02832) | 0.186 | 0.566 |
| LV mass | Hippocampal volume | 964 | 2,848 | 237 / 727 | −0.00341 (−0.00888 to 0.00206) | 0.222 | 0.566 |
| LA dimension | Hippocampal volume | 1,305 | 3,787 | 303 / 1,002 | −0.00274 (−0.00739 to 0.00191) | 0.248 | 0.566 |
| LV mass | Total brain volume | 964 | 2,848 | 237 / 727 | −0.00202 (−0.00593 to 0.00189) | 0.311 | 0.575 |
| LV diastolic dimension | Lateral ventricular volume | 983 | 2,904 | 241 / 742 | −0.00281 (−0.00864 to 0.00302) | 0.345 | 0.575 |
| LV mass | Lateral ventricular volume | 964 | 2,848 | 237 / 727 | −0.00244 (−0.00769 to 0.00281) | 0.362 | 0.575 |
| LV diastolic dimension | White matter hyperintensity burden | 650 | 1,442 | 158 / 492 | 0.00786 (−0.01025 to 0.02597) | 0.395 | 0.575 |
| Fractional shortening | Hippocampal volume | 952 | 2,812 | 235 / 717 | 0.00168 (−0.00318 to 0.00653) | 0.498 | 0.664 |
| LV diastolic dimension | Hippocampal volume | 983 | 2,904 | 241 / 742 | −0.00162 (−0.00763 to 0.00439) | 0.597 | 0.712 |
| LA dimension | White matter hyperintensity burden | 809 | 1,792 | 185 / 624 | −0.00370 (−0.01845 to 0.01105) | 0.623 | 0.712 |
| LA dimension | Total brain volume | 1,305 | 3,787 | 303 / 1,002 | 0.00066 (−0.00262 to 0.00394) | 0.694 | 0.741 |
| LV diastolic dimension | Total brain volume | 983 | 2,904 | 241 / 742 | 0.00001 (−0.00427 to 0.00430) | 1.00 | 1.00 |

Panel B. The same 16 tests refitted with participant-specific random intercepts and MRI-time slopes, with age × MRI time and sex × MRI time terms; FDR correction across the 16 tests.

| Cardiac trajectory | Brain outcome | N subjects | MRI observations | β, random-slope model (95% CI) | P | FDR q |
|---|---|---|---|---|---|---|
| LA dimension | Lateral ventricular volume | 1,305 | 3,787 | −0.00883 (−0.01445 to −0.00320) | 0.0021 | 0.0336 |
| Fractional shortening | Lateral ventricular volume | 952 | 2,812 | 0.00498 (−0.00112 to 0.01108) | 0.11 | 0.592 |
| Fractional shortening | Total brain volume | 952 | 2,812 | 0.00248 (−0.00095 to 0.00591) | 0.156 | 0.592 |
| Fractional shortening | White matter hyperintensity volume | 630 | 1,395 | −0.01109 (−0.02692 to 0.00474) | 0.17 | 0.592 |
| LV end-diastolic dimension | White matter hyperintensity volume | 650 | 1,442 | 0.01241 (−0.00708 to 0.03189) | 0.212 | 0.592 |
| LV mass | Lateral ventricular volume | 964 | 2,848 | −0.00433 (−0.01134 to 0.00268) | 0.226 | 0.592 |
| LV mass | Total brain volume | 964 | 2,848 | −0.00224 (−0.00612 to 0.00165) | 0.259 | 0.592 |
| Fractional shortening | Hippocampal volume | 952 | 2,812 | 0.00263 (−0.00279 to 0.00806) | 0.341 | 0.683 |
| LV mass | White matter hyperintensity volume | 638 | 1,415 | 0.00751 (−0.01115 to 0.02618) | 0.43 | 0.765 |
| LV mass | Hippocampal volume | 964 | 2,848 | −0.00212 (−0.00826 to 0.00402) | 0.499 | 0.799 |
| LA dimension | Hippocampal volume | 1,305 | 3,787 | −0.00118 (−0.00646 to 0.00409) | 0.66 | 0.89 |
| LA dimension | Total brain volume | 1,305 | 3,787 | 0.00071 (−0.00254 to 0.00396) | 0.667 | 0.89 |
| LV end-diastolic dimension | Lateral ventricular volume | 983 | 2,904 | −0.00106 (−0.00855 to 0.00642) | 0.781 | 0.926 |
| LV end-diastolic dimension | Total brain volume | 983 | 2,904 | −0.00078 (−0.00715 to 0.00559) | 0.81 | 0.926 |
| LA dimension | White matter hyperintensity volume | 809 | 1,792 | 0.00100 (−0.01550 to 0.01751) | 0.905 | 0.965 |
| LV end-diastolic dimension | Hippocampal volume | 983 | 2,904 | 0.00027 (−0.07834 to 0.07888) | 0.995 | 0.995 |

All 16 cardiac trajectory × brain outcome combinations in the prospective sample, ordered by P. Each model used a participant-specific random intercept and adjusted for age at Examination 6 and sex; cardiac trajectories and brain outcomes were standardized within each analytic sample. FDR q values are Benjamini–Hochberg adjusted across the 16 tests. Sample sizes differ by availability of the cardiac trajectory and brain outcome.

Abbreviations: APOE, apolipoprotein E; CI, confidence interval; FDR, false discovery rate; LA, left atrial; LV, left ventricular.

#### Table S3. Sensitivity of the LA remodeling × MRI time × APOE ε4 association to longitudinal model specification, exposure definition, head size, and early vascular covariates

| Model / sensitivity analysis | N | MRI observations | Interaction β (95% CI) | P |
|---|---|---|---|---|
| Primary random-intercept model | 1,305 | 3,787 | −0.00858 (−0.01300 to −0.00415) | $1.45 \times 10^{-4}$ |
| + age × MRI time + sex × MRI time | 1,305 | 3,787 | −0.00877 (−0.01268 to −0.00485) | $1.14 \times 10^{-5}$ |
| + subject-specific random MRI-time slope | 1,305 | 3,787 | −0.00883 (−0.01445 to −0.00320) | 0.00210 |
| + baseline LA dimension + baseline LA × MRI time | 1,305 | 3,787 | −0.00880 (−0.01442 to −0.00317) | 0.00223 |
| Examination 4→6 annualized LA change | 1,356 | 3,924 | −0.00759 (−0.01305 to −0.00212) | 0.00650 |
| ICV-complete matched sample, no ICV terms | 795 | 2,405 | −0.00451 (−0.00813 to −0.00090) | 0.0144 |
| + ICV | 795 | 2,405 | −0.00449 (−0.00811 to −0.00088) | 0.0148 |
| + ICV + ICV × MRI time | 795 | 2,405 | −0.00334 (−0.00680 to 0.00011) | 0.0581 |
| Early vascular covariates | 1,269 | 3,685 | −0.00882 (−0.01333 to −0.00431) | $1.25 \times 10^{-4}$ |
| Early vascular covariates + DBP | 1,269 | 3,685 | −0.00882 (−0.01333 to −0.00431) | $1.26 \times 10^{-4}$ |
| Matched sample with Exam 4 antihypertensive data | 1,268 | 3,682 | −0.00890 (−0.01341 to −0.00439) | $1.09 \times 10^{-4}$ |
| + Exam 4 antihypertensive treatment | 1,268 | 3,682 | −0.00893 (−0.01344 to −0.00442) | $1.05 \times 10^{-4}$ |

Rows 2–4 are cumulative: each adds terms to the model in the row above. The baseline-LA model therefore includes age × MRI time, sex × MRI time, a participant-specific random MRI-time slope, baseline LA dimension, and baseline LA dimension × MRI time. This expanded model was used for the native-unit interpretation shown in Figure 2A.

The Examination 4→6 analysis defined LA remodeling as the change in LA dimension divided by the elapsed time between dated Examinations 4 and 6, with Examination 4 LA dimension and its interaction with MRI time included.

Head-size analyses were restricted to participants with available intracranial volume (ICV) and fitted with a random intercept. Comparing the no-ICV row with the principal model shows the effect of sample restriction; comparing the ICV rows with the no-ICV row shows the effect of ICV adjustment within the same sample. A random-slope ICV specification did not converge and is not reported.

Early vascular covariates: Examination 4 body mass index, systolic blood pressure, smoking, and diabetes, added to the principal random-intercept model; the last row additionally includes diastolic blood pressure. N is lower than in the principal model because of missing covariates.

Coefficients are in standardized brain-outcome units per year per SD of LA remodeling. Abbreviations: APOE, apolipoprotein E; CI, confidence interval; DBP, diastolic blood pressure; ICV, intracranial volume; LA, left atrial.

#### Table S4. Availability of subsequent brain MRI in the pre-MRI at-risk cohort, and selection into the repeated-MRI cohort with inverse-probability-of-selection weighting

***Panel A. Overall (N=2,522)***

| Eligible subsequent MRI examinations | n | % |
|---|---|---|
| ≥2 eligible MRI | 1,305 | 51.7 |
| 1 eligible MRI | 339 | 13.4 |
| 0 eligible MRI | 878 | 34.8 |

***Panel B. By APOE ε4 status and tertile of antecedent LA remodeling***

| APOE ε4 status | LA remodeling tertile | N | ≥2 eligible MRI, n (%) | 1 eligible MRI, n (%) | 0 eligible MRI, n (%) |
|---|---|---|---|---|---|
| APOE ε4 carrier | T1: lower remodeling | 181 | 93 (51.4%) | 12 (6.6%) | 76 (42.0%) |
| APOE ε4 carrier | T2: middle | 188 | 114 (60.6%) | 20 (10.6%) | 54 (28.7%) |
| APOE ε4 carrier | T3: greater remodeling | 186 | 96 (51.6%) | 27 (14.5%) | 63 (33.9%) |
| APOE ε4 noncarrier | T1: lower remodeling | 660 | 336 (50.9%) | 98 (14.8%) | 226 (34.2%) |
| APOE ε4 noncarrier | T2: middle | 652 | 347 (53.2%) | 91 (14.0%) | 214 (32.8%) |
| APOE ε4 noncarrier | T3: greater remodeling | 655 | 319 (48.7%) | 91 (13.9%) | 245 (37.4%) |

At-risk cohort: participants with a valid three-examination LA remodeling trajectory (Examinations 4–6) and classified APOE genotype, defined before conditioning on subsequent MRI participation. Inclusion in the principal longitudinal analysis required at least two eligible lateral ventricular MRI observations after the final cardiac examination. Tertiles of LA remodeling were defined in the at-risk cohort. Percentages are row percentages.

Abbreviations: APOE, apolipoprotein E; LA, left atrial.

#### Selection model and inverse-probability-of-selection weighted sensitivity analysis

***Panel A. Selection into the repeated-MRI cohort***

| Quantity | N | Estimate (95% CI) | P |
| --- | --- | --- | --- |
| At-risk cohort | 2,522 | — | — |
| Selection-model complete cases (included / not included) | 2,347 (1,269 / 1,078) | — | — |
| LA remodeling × APOE ε4 association with inclusion, OR | 2,347 | 1.065 (0.880 to 1.288) | 0.519 |
| Stabilized weight, 1st / 99th percentile (truncation limits) | 2,347 | 0.690 / 1.580 | — |

***Panel B. LA remodeling × MRI time × APOE ε4 interaction with and without selection weighting***

| Model | N | MRI observations | Interaction β (95% CI) | P |
| --- | --- | --- | --- | --- |
| Primary mixed model | 1,305 | 3,787 | −0.00858 (−0.01300 to −0.00415) | $1.45\times10^{-4}$ |
| Unweighted GEE, IPSW-complete sample | 1,269 | 3,685 | −0.00869 (−0.01683 to −0.00056) | 0.0361 |
| IPSW-weighted GEE, weights truncated at 1st/99th percentiles | 1,269 | 3,685 | −0.00839 (−0.01630 to −0.00049) | 0.0373 |

Selection model: logistic regression of inclusion on standardized LA remodeling, APOE ε4 carrier status, their interaction, age at Examination 6, sex, and Examination 4 body mass index, systolic blood pressure, diabetes, smoking, and LA dimension. Stabilized weights were P(inclusion | APOE ε4)/P(inclusion | full covariate model), truncated at the 1st and 99th percentiles.

Weighted and unweighted GEE used an exchangeable working correlation and robust standard errors and were fitted to the same complete-case participants; the comparison between them is the like-for-like test of selection. The principal mixed-model estimate is shown for reference.

Abbreviations: APOE, apolipoprotein E; CI, confidence interval; GEE, generalized estimating equations; IPSW, inverse-probability-of-selection weighting; LA, left atrial; OR, odds ratio.

#### Table S5. Antecedent cardiac trajectory × APOE ε4 interactions with later plasma biomarkers and later cognition

| Cardiac trajectory | Plasma biomarker | N | Noncarriers / carriers, n | Interaction β (95% CI) | P | q (within trajectory, 4 tests) | q (global, 16 tests) |
|---|---|---|---|---|---|---|---|
| LA dimension | Aβ42/40 | 1,620 | 1,266 / 354 | −0.064 (−0.1624 to 0.0352) | 0.207 | 0.451 | 0.740 |
| LA dimension | p-tau181 | 1,308 | 1,019 / 289 | −0.068 (−0.1784 to 0.0421) | 0.226 | 0.451 | 0.740 |
| LA dimension | GFAP | 1,313 | 1,022 / 291 | 0.007 (−0.1037 to 0.1177) | 0.901 | 0.901 | 0.976 |
| LA dimension | NfL | 1,313 | 1,022 / 291 | 0.023 (−0.0705 to 0.1165) | 0.630 | 0.840 | 0.916 |
| LV mass | Aβ42/40 | 1,127 | 870 / 257 | 0.073 (−0.0502 to 0.1965) | 0.245 | 0.380 | 0.740 |
| LV mass | p-tau181 | 943 | 724 / 219 | −0.003 (−0.1719 to 0.1654) | 0.970 | 0.970 | 0.976 |
| LV mass | GFAP | 946 | 725 / 221 | −0.073 (−0.2079 to 0.0611) | 0.285 | 0.380 | 0.740 |
| LV mass | NfL | 946 | 725 / 221 | −0.083 (−0.2351 to 0.0682) | 0.281 | 0.380 | 0.740 |
| Fractional shortening | Aβ42/40 | 1,118 | 862 / 256 | −0.050 (−0.1816 to 0.0818) | 0.458 | 0.611 | 0.740 |
| Fractional shortening | p-tau181 | 930 | 714 / 216 | 0.062 (−0.0548 to 0.1796) | 0.297 | 0.611 | 0.740 |
| Fractional shortening | GFAP | 933 | 715 / 218 | 0.063 (−0.0645 to 0.1903) | 0.333 | 0.611 | 0.740 |
| Fractional shortening | NfL | 933 | 715 / 218 | 0.011 (−0.1009 to 0.1223) | 0.851 | 0.851 | 0.976 |
| LV end-diastolic dimension | Aβ42/40 | 1,149 | 887 / 262 | 0.050 (−0.0841 to 0.1850) | 0.462 | 0.925 | 0.740 |
| LV end-diastolic dimension | p-tau181 | 957 | 736 / 221 | 0.072 (−0.0859 to 0.2300) | 0.371 | 0.925 | 0.740 |
| LV end-diastolic dimension | GFAP | 960 | 737 / 223 | 0.014 (−0.1183 to 0.1461) | 0.837 | 0.976 | 0.976 |
| LV end-diastolic dimension | NfL | 960 | 737 / 223 | 0.002 (−0.1344 to 0.1385) | 0.976 | 0.976 | 0.976 |

Exposures: participant-specific slopes of echocardiographic measures across Offspring Examinations 4–6. Outcomes: later plasma biomarkers. FDR q values are reported within each cardiac trajectory (4 tests) and across all 16 tests (global). No interaction met global q<0.05.

This analysis is distinct from the biomarker adjustment of the regional cardiac–brain interactions (Figure S2) and from the cross-sectional cardiac phenotype × APOE ε4 associations with plasma biomarkers in the regional cohort).

Abbreviations: Aβ, amyloid-β; APOE, apolipoprotein E; CI, confidence interval; GFAP, glial fibrillary acidic protein; LV, left ventricular; NfL, neurofilament light chain; p-tau181, tau phosphorylated at threonine 181.

#### Later cognition

***Panel A. Later neuropsychological tests***

| Cardiac trajectory | Cognitive test | N | Interaction β (95% CI) | P | q (global, 16 tests) | q (within trajectory, 4 tests) |
|---|---|---|---|---|---|---|
| LA dimension | Logical Memory delayed | 364 | 0.154 (−0.1347 to 0.4422) | 0.296 | 0.730 | 0.596 |
| LA dimension | Trails B | 279 | 0.008 (−0.3589 to 0.3741) | 0.968 | 0.968 | 0.968 |
| LA dimension | Animal fluency | 356 | 0.120 (−0.1062 to 0.3464) | 0.298 | 0.730 | 0.596 |
| LA dimension | Digit Span Backward | 346 | 0.051 (−0.1994 to 0.3005) | 0.692 | 0.923 | 0.923 |
| LV mass | Logical Memory delayed | 304 | −0.201 (−0.5354 to 0.1330) | 0.238 | 0.730 | 0.426 |
| LV mass | Trails B | 232 | −0.185 (−0.5484 to 0.1788) | 0.319 | 0.730 | 0.426 |
| LV mass | Animal fluency | 297 | 0.025 (−0.2498 to 0.2992) | 0.860 | 0.968 | 0.860 |
| LV mass | Digit Span Backward | 292 | −0.198 (−0.4689 to 0.0735) | 0.153 | 0.730 | 0.426 |
| Fractional shortening | Logical Memory delayed | 300 | 0.018 (−0.3034 to 0.3389) | 0.914 | 0.968 | 0.914 |
| Fractional shortening | Trails B | 229 | 0.063 (−0.2126 to 0.3377) | 0.656 | 0.923 | 0.874 |
| Fractional shortening | Animal fluency | 293 | 0.136 (−0.2498 to 0.5226) | 0.489 | 0.869 | 0.874 |
| Fractional shortening | Digit Span Backward | 288 | −0.083 (−0.3110 to 0.1454) | 0.477 | 0.869 | 0.874 |
| LV end-diastolic dimension | Logical Memory delayed | 308 | −0.246 (−0.6710 to 0.1798) | 0.258 | 0.730 | 0.516 |
| LV end-diastolic dimension | Trails B | 235 | −0.178 (−0.8277 to 0.4724) | 0.592 | 0.923 | 0.790 |
| LV end-diastolic dimension | Animal fluency | 301 | −0.059 (−0.5094 to 0.3916) | 0.798 | 0.968 | 0.798 |
| LV end-diastolic dimension | Digit Span Backward | 296 | −0.472 (−0.8363 to −0.1072) | 0.0112 | 0.179 | 0.0448 |

***Panel B. Repeated MMSE: cardiac trajectory × cognitive time × APOE ε4***

| Cardiac trajectory | N | MMSE observations | Interaction β (95% CI) | P | FDR q (4 tests) |
|---|---|---|---|---|---|
| LA dimension | 1,670 | 8,730 | −0.00159 (−0.0066 to 0.0034) | 0.531 | 0.708 |
| LV mass | 1,205 | 6,409 | −0.00786 (−0.0136 to −0.0021) | 0.00731 | 0.0292 |
| Fractional shortening | 1,193 | 6,340 | 0.00182 (−0.0034 to 0.0070) | 0.492 | 0.708 |
| LV end-diastolic dimension | 1,229 | 6,531 | −0.00041 (−0.0065 to 0.0057) | 0.897 | 0.897 |

Panel A: later neuropsychological test scores at the first eligible later examination (Logical Memory delayed recall, Trails B, animal fluency, Digit Span Backward); the primary correction family is the global 16 tests, with within-trajectory q values shown for reference. The LV end-diastolic dimension × APOE ε4 interaction for Digit Span Backward has within-trajectory q=0.045 but global q=0.179 and is not considered significant.

Panel B: repeated MMSE-based scores in participants with at least two eligible post-exposure administrations; FDR across the four cardiac trajectories. The ≥2-year-gap analysis gave the same inference (archived with the source data).

Abbreviations: APOE, apolipoprotein E; CI, confidence interval; FDR, false discovery rate; LA, left atrial; LV, left ventricular; MMSE, Mini-Mental State Examination.

**Table S6. Regional cardiac–brain associations meeting the FDR criterion for the cardiac main effect or the cardiac × APOE ε4 interaction**

| Model | Anatomical family | Brain phenotype | Cardiac phenotype | N | β cardiac | P cardiac | q cardiac | β interaction (95% CI) | P interaction | q interaction |
|---|---|---|---|---|---|---|---|---|---|---|
| M1 | Cingulate | Left S cingul Marginalis surface area | LVEDVi | 782 | −0.128 | $3.16 \times 10^{-4}$ | 0.00631 | — | — | — |
| M1 | Cingulate | Left S cingul Marginalis volume | LVESVi | 782 | −0.128 | $4.76 \times 10^{-4}$ | 0.00953 | — | — | — |
| M1 | Cingulate | Left S cingul Marginalis surface area | LVESVi | 782 | −0.124 | $6.81 \times 10^{-4}$ | 0.0136 | — | — | — |
| M1 | Other temporal | Left transversetemporal volume | LVEF | 785 | 0.109 | 0.00112 | 0.0224 | — | — | — |
| M1 | Occipital | Left G occipital sup surface area | LV mass index | 782 | −0.117 | 0.00120 | 0.0261 | — | — | — |
| M1 | Occipital | Left S oc temp med Lingual surface area | LV mass index | 782 | −0.112 | 0.00191 | 0.0261 | — | — | — |
| M1 | Occipital | Left lingual surface area | LV mass index | 782 | −0.115 | 0.00280 | 0.0261 | — | — | — |
| M1 | Medial temporal/AD | Left Pole temporal surface area | LVESVi | 782 | −0.093 | 0.00337 | 0.0270 | — | — | — |
| M1 | Cingulate | Left S cingul Marginalis volume | LVEDVi | 782 | −0.112 | 0.00336 | 0.0336 | — | — | — |
| M1 | Cingulate | Right rostralanteriorcingulate volume | LVEDVi | 782 | 0.117 | 0.00213 | 0.0336 | — | — | — |
| M1 | Frontal | Left G precentral volume | LVEF | 785 | 0.091 | 0.00437 | 0.0481 | — | — | — |
| M1 | Frontal | Left precentral volume | LVEF | 785 | 0.098 | 0.00335 | 0.0481 | — | — | — |
| M2 | Other temporal | Left transversetemporal volume | LVEF | 781 | 0.112 | $9.34 \times 10^{-4}$ | 0.0187 | — | — | — |
| M2 | Cingulate | Left S cingul Marginalis volume | LVESVi | 781 | −0.124 | 0.00108 | 0.0217 | — | — | — |
| M2 | Occipital | Left G occipital sup surface area | LV mass index | 781 | −0.127 | 0.00101 | 0.0235 | — | — | — |
| M2 | Occipital | Left lingual surface area | LV mass index | 781 | −0.127 | 0.00168 | 0.0235 | — | — | — |
| M2 | Cingulate | Left S cingul Marginalis surface area | LVEDVi | 781 | −0.117 | 0.00163 | 0.0327 | — | — | — |
| M2 | Occipital | Left S oc temp med Lingual surface area | LV mass index | 781 | −0.110 | 0.00385 | 0.0359 | — | — | — |
| M2 | Frontal | Left G precentral volume | LVEF | 781 | 0.094 | 0.00367 | 0.0404 | — | — | — |
| M2 | Frontal | Left precentral volume | LVEF | 781 | 0.101 | 0.00244 | 0.0404 | — | — | — |
| M2 | Medial temporal/AD | Left Pole temporal surface area | LVESVi | 781 | −0.091 | 0.00581 | 0.0465 | — | — | — |
| M3 | Frontal | Right superior frontal surface area | LVESVi | 765 | 0.035 | 0.142 | 0.926 | −0.170 (−0.260 to −0.081) | $1.97 \times 10^{-4}$ | 0.00433 |
| M3 | Occipital | Right lingual volume | LV mass index | 765 | −0.092 | 0.0517 | 0.362 | 0.305 (0.144 to 0.467) | $2.16 \times 10^{-4}$ | 0.00604 |
| M3 | Occipital | Right lingual surface area | LV mass index | 765 | −0.106 | 0.00772 | 0.0720 | 0.226 (0.095 to 0.357) | $7.09 \times 10^{-4}$ | 0.0198 |
| M3 | Other temporal | Left middle temporal volume | LVEF | 765 | −0.046 | 0.234 | 0.511 | 0.233 (0.087 to 0.380) | 0.00176 | 0.0352 |
| M3 | Medial temporal/AD | Left Pole temporal surface area | LVEDVi | 765 | −0.103 | 0.00525 | 0.0420 | 0.130 (−0.020 to 0.280) | 0.0901 | 0.240 |
| M3 | Medial temporal/AD | Left Pole temporal surface area | LVESVi | 765 | −0.113 | 0.00133 | 0.0106 | 0.098 (−0.071 to 0.267) | 0.257 | 0.586 |

| Model | Anatomical family | Brain phenotype | Cardiac phenotype | N | β cardiac | P cardiac | q cardiac | β interaction (95% CI) | P interaction | q interaction |
|---|---|---|---|---|---|---|---|---|---|---|
| M3 | Cingulate | Right isthmuscingulate surface area | LV mass index | 765 | −0.121 | 0.00222 | 0.0443 | 0.051 (−0.079 to 0.182) | 0.443 | 0.859 |

M1: age at brain MRI, sex, and absolute CMR–brain MRI interval. M2: M1 plus body mass index, systolic blood pressure, current smoking, and diabetes history. M3: M2 plus APOE ε4 carrier status and the cardiac phenotype × APOE ε4 interaction; in M3, β cardiac is the slope among APOE ε4 noncarriers. Surface-area models additionally adjusted for total cortical surface area; regional volumes were divided by intracranial volume. Continuous variables were standardized within model-specific complete cases; HC3 robust standard errors were used.

q values are Benjamini–Hochberg adjusted within each brain metric × anatomical family × cardiac phenotype testing group. Rows are shown if $q<0.05$ for the cardiac main effect or the interaction. Destrieux parcels retain atlas abbreviations (G, gyrus; S, sulcus).

Complete results for all M1, M2, and M3 models (1,960 tests per model) are provided in Supplementary Data S1 (Excel workbook).

Abbreviations: AD, Alzheimer disease; APOE, apolipoprotein E; CI, confidence interval; CMR, cardiac magnetic resonance; FDR, false discovery rate; LVEDVi, LV end-diastolic volume index; LVEF, LV ejection fraction; LVESVi, LV end-systolic volume index.

##### Table S7. Sensitivity of the four regional ventricular cardiac × APOE ε4 interactions to metabolic markers, and pedigree-family cluster–robust inference

*Panel A. Individual metabolic markers, matched complete cases*

| Association | Marker | N | Noncarriers / carriers, n | Matched M3 interaction β | Interaction β after adjustment | Change in \|β\|, % | P after adjustment |
|---|---|---|---|---|---|---|---|
| LVEF × APOE ε4 → left middle temporal volume | HbA1c | 653 | 502 / 151 | 0.186 | 0.183 | 1.7 | 0.0145 |
| LVEF × APOE ε4 → left middle temporal volume | Insulin | 764 | 592 / 172 | 0.202 | 0.198 | 1.6 | 0.0056 |
| LVEF × APOE ε4 → left middle temporal volume | Adiponectin | 601 | 462 / 139 | 0.187 | 0.183 | 1.9 | 0.0182 |
| LVESVi × APOE ε4 → right superior frontal surface area | HbA1c | 653 | 502 / 151 | −0.180 | −0.179 | 0.3 | 0.0002 |
| LVESVi × APOE ε4 → right superior frontal surface area | Insulin | 764 | 592 / 172 | −0.172 | −0.169 | 1.8 | 0.0002 |
| LVESVi × APOE ε4 → right superior frontal surface area | Adiponectin | 601 | 462 / 139 | −0.187 | −0.183 | 2.1 | 0.0003 |
| LV mass index × APOE ε4 → right lingual volume | HbA1c | 653 | 502 / 151 | 0.259 | 0.260 | −0.2 | 0.0018 |
| LV mass index × APOE ε4 → right lingual volume | Insulin | 764 | 592 / 172 | 0.250 | 0.255 | −2.0 | 0.0008 |
| LV mass index × APOE ε4 → right lingual volume | Adiponectin | 601 | 462 / 139 | 0.220 | 0.222 | −0.7 | 0.0090 |
| LV mass index × APOE ε4 → right lingual surface area | HbA1c | 653 | 502 / 151 | 0.220 | 0.220 | 0.0 | 0.0026 |
| LV mass index × APOE ε4 → right lingual surface area | Insulin | 764 | 592 / 172 | 0.228 | 0.225 | 1.3 | 0.0007 |
| LV mass index × APOE ε4 → right lingual surface area | Adiponectin | 601 | 462 / 139 | 0.183 | 0.181 | 1.2 | 0.0170 |

*Panel B. Joint adjustment for HbA1c, insulin, and adiponectin (common complete-case sample)*

| Association | N | Noncarriers / carriers, n | Matched M3 β (95% CI) | P | Adjusted β (95% CI) | P | Change in \|β\|, % |
|---|---|---|---|---|---|---|---|
| LVEF × APOE ε4 → left middle temporal volume | 600 | 461 / 139 | 0.186 (0.034 to 0.338) | 0.0163 | 0.173 (0.023 to 0.323) | 0.0240 | 7.2 |
| LVESVi × APOE ε4 → right superior frontal surface area | 600 | 461 / 139 | −0.187 (−0.286 to −0.088) | 0.0002 | −0.180 (−0.275 to −0.085) | 0.0002 | 4.1 |
| LV mass index × APOE ε4 → right lingual volume | 600 | 461 / 139 | 0.224 (0.057 to 0.390) | 0.0084 | 0.227 (0.061 to 0.394) | 0.0074 | −1.7 |
| LV mass index × APOE ε4 → right lingual surface area | 600 | 461 / 139 | 0.186 (0.038 to 0.334) | 0.0141 | 0.177 (0.028 to 0.326) | 0.0197 | 4.7 |

*Panel C. Cardiac × APOE ε4 × metabolic marker three-way interactions*

| Association | Marker | N | Noncarriers / carriers, n | Three-way β (95% CI) | P | FDR q |
|---|---|---|---|---|---|---|
| LV mass index × APOE ε4 → right lingual volume | Insulin | 764 | 592 / 172 | −0.173 (−0.348 to 0.003) | 0.0541 | 0.4804 |

| Association | Marker | N | Noncarriers / carriers, n | Three-way β (95% CI) | P | FDR q |
|---|---|---|---|---|---|---|
| LVEF × APOE ε4 → left middle temporal volume | Insulin | 764 | 592 / 172 | −0.112 (−0.257 to 0.033) | 0.1314 | 0.4804 |
| LVESVi × APOE ε4 → right superior frontal surface area | HbA1c | 653 | 502 / 151 | 0.099 (−0.033 to 0.231) | 0.1429 | 0.4804 |
| LVESVi × APOE ε4 → right superior frontal surface area | Adiponectin | 601 | 462 / 139 | 0.073 (−0.029 to 0.175) | 0.1601 | 0.4804 |
| LVEF × APOE ε4 → left middle temporal volume | HbA1c | 653 | 502 / 151 | 0.176 (−0.096 to 0.448) | 0.2043 | 0.4903 |
| LV mass index × APOE ε4 → right lingual volume | HbA1c | 653 | 502 / 151 | 0.110 (−0.111 to 0.331) | 0.3302 | 0.4925 |
| LVESVi × APOE ε4 → right superior frontal surface area | Insulin | 764 | 592 / 172 | −0.043 (−0.136 to 0.049) | 0.3575 | 0.4925 |
| LV mass index × APOE ε4 → right lingual surface area | Insulin | 764 | 592 / 172 | −0.070 (−0.224 to 0.084) | 0.3725 | 0.4925 |
| LV mass index × APOE ε4 → right lingual volume | Adiponectin | 601 | 462 / 139 | −0.072 (−0.239 to 0.095) | 0.3985 | 0.4925 |
| LV mass index × APOE ε4 → right lingual surface area | Adiponectin | 601 | 462 / 139 | −0.055 (−0.195 to 0.084) | 0.4380 | 0.4925 |
| LVEF × APOE ε4 → left middle temporal volume | Adiponectin | 601 | 462 / 139 | 0.062 (−0.102 to 0.227) | 0.4571 | 0.4925 |
| LV mass index × APOE ε4 → right lingual surface area | HbA1c | 653 | 502 / 151 | 0.056 (−0.104 to 0.216) | 0.4925 | 0.4925 |

Models used the M3 covariates (age, sex, absolute CMR–brain MRI interval, body mass index, systolic blood pressure, current smoking, diabetes history, and total cortical surface area for surface-area outcomes) with HC3 robust standard errors. Outcomes and cardiac phenotypes were standardized. Insulin and adiponectin were log-transformed before standardization.

Panels A and B compare each adjusted model with the M3 model fitted in the same participants, so differences reflect adjustment rather than sample membership. Change in |β| is relative to the matched M3 estimate; negative values indicate a slightly larger absolute interaction after adjustment. Panel C: FDR correction across 12 prespecified three-way tests (4 associations × 3 markers). These analyses assess attenuation and do not test mediation.

Abbreviations: APOE, apolipoprotein E; CI, confidence interval; FDR, false discovery rate; HbA1c, glycated hemoglobin; LVEF, LV ejection fraction; LVESVi, LV end-systolic volume index.

#### Pedigree-family cluster–robust inference for the cardiac × APOE ε4 interactions and the exploratory HbA1c × APOE ε4 interaction

| Analysis | N | Pedigrees | Interaction β (95% CI) | HC3 SE | Cluster-robust SE | Cluster/HC3 SE ratio | Cluster-robust P |
|---|---|---|---|---|---|---|---|
| LVEF × APOE ε4 → left middle temporal volume | 724 | 377 | 0.212 (0.076 to 0.348) | 0.077 | 0.069 | 0.90 | 0.00225 |
| LVESVi × APOE ε4 → right superior frontal surface area | 724 | 377 | −0.134 (−0.224 to −0.043) | 0.048 | 0.046 | 0.96 | 0.00368 |
| LV mass index × APOE ε4 → right lingual volume | 724 | 377 | 0.319 (0.173 to 0.465) | 0.085 | 0.074 | 0.88 | $1.79 \times 10^{-5}$ |
| LV mass index × APOE ε4 → right lingual surface area | 724 | 377 | 0.210 (0.083 to 0.337) | 0.068 | 0.065 | 0.96 | 0.00119 |
| LAVI × APOE ε4 → left middle-occipital cortical-thickness variability | 433 | 271 | −0.582 (−0.796 to −0.367) | 0.121 | 0.109 | 0.90 | $1.06 \times 10^{-7}$ |
| LVEDVi × APOE ε4 × HbA1c | 612 | 344 | −2.85 (−4.99 to −0.71) | 1.20 | 1.09 | 0.91 | 0.00908 |

Models were refitted in participants with pedigree information, with standard errors clustered on Framingham pedigree family number. HC3 standard errors were computed in the same samples for comparison; 95% CIs and P values are cluster-robust. Ventricular models used the fully adjusted M3 specification (standardized cardiac phenotype, APOE ε4 carrier status, their interaction, age at brain MRI, sex, absolute CMR–brain MRI interval, body mass index, systolic blood pressure, current smoking, and diabetes history); regional volumes were divided by intracranial volume before standardization, and surface-area models adjusted for total cortical surface area. The atrial model used the corresponding atrial covariates. The HbA1c model used the specification of Table S8 (age at CMR, sex, HbA1c-to-CMR interval, body mass index, systolic blood pressure, and antihypertensive medication); outcome LVEDVi in mL/m² per SD of HbA1c.

Among the 724 participants in the ventricular analyses, 513 belonged to 166 pedigrees represented by more than one analyzed participant; the remaining 211 pedigrees were represented by one participant each.

Continuous variables in the heart–brain models were standardized within each pedigree-complete sample, so coefficients are on the scale of the main tables. HC3 and cluster-robust models fitted to the same sample give identical coefficients; differences from the primary estimates (Table 2, Table 4) therefore reflect the smaller pedigree-complete samples rather than the variance estimator.

Pedigree-family cluster–robust inference accounts for within-family dependence but is not equivalent to a genetic kinship matrix or pedigree random-effects model.

Abbreviations: APOE, apolipoprotein E; CI, confidence interval; CMR, cardiac magnetic resonance; HbA1c, glycated hemoglobin; LAVI, left atrial volume index (maximum); LVEDVi, LV end-diastolic volume index; LVEF, LV ejection fraction; LVESVi, LV end-systolic volume index; SE, standard error.

#### Table S8. APOE ε4 modification of the association between HbA1c and LV end-diastolic volume index

| Model | N | HbA1c × APOE ε4 β (95% CI) | P | Noncarrier slope (95% CI) | P | Carrier slope (95% CI) | P | $R^2$ |
|---|---|---|---|---|---|---|---|---|
| M0: Age + sex | 653 | −2.93 (−5.09 to −0.77) | 0.00783 | −0.33 (−1.62 to 0.95) | 0.609 | −3.26 (−5.00 to −1.52) | $2.38 \times 10^{-4}$ | 0.151 |
| M1: + HbA1c-CMR interval | 653 | −2.86 (−4.99 to −0.73) | 0.00858 | −0.34 (−1.64 to 0.97) | 0.613 | −3.20 (−4.88 to −1.51) | $2.06 \times 10^{-4}$ | 0.159 |
| M2: + BMI | 653 | −2.79 (−5.02 to −0.56) | 0.0143 | −0.18 (−1.49 to 1.13) | 0.786 | −2.97 (−4.79 to −1.15) | 0.00135 | 0.182 |
| M3: + SBP + antihypertensive medication | 650 | −2.89 (−5.21 to −0.56) | 0.0148 | −0.17 (−1.55 to 1.20) | 0.804 | −3.06 (−4.94 to −1.18) | 0.00141 | 0.187 |

Outcome: LVEDVi (mL/m²). HbA1c was standardized; coefficients are mL/m² per 1-SD higher HbA1c. Models are nested (each adds covariates to the row above). HC3 robust standard errors. This analysis followed the single FDR-significant result of the exploratory discovery analysis (Table S9).

Abbreviations: APOE, apolipoprotein E; CI, confidence interval; CMR, cardiac magnetic resonance; HbA1c, glycated hemoglobin; LVEDVi, LV end-diastolic volume index; SBP, systolic blood pressure.

##### Table S9. Exploratory APOE ε4 × metabolic-modifier analyses across brain and cardiac phenotype families

| Modifier | Domain | Phenotype family | Tests, n | FDR q<0.05, n | Nominal P<0.05, n | Top-ranked phenotype | Interaction β | P | FDR q |
|---|---|---|---|---|---|---|---|---|---|
| Current diabetes | Brain | Area | 220 | 0 | 7 | Right frontalpole surface area | −0.777 | 0.00789 | 0.991 |
| Current diabetes | Brain | Mean curvature | 216 | 0 | 4 | Left G front inf Opercular mean curvature | −0.650 | 0.0140 | 0.983 |
| Current diabetes | Brain | Cortical thickness | 216 | 0 | 2 | Right S oc middle and Lunatus thickness | −0.512 | 0.0289 | 0.950 |
| Current diabetes | Brain | Thickness variability | 216 | 0 | 5 | Right Pole occipital thickness SD | 0.727 | 0.0175 | 0.917 |
| Current diabetes | Brain | Volume | 216 | 0 | 9 | Left G pariet inf Supramar volume | −0.613 | 0.00949 | 0.920 |
| Current diabetes | Heart | Atrial | 3 | 0 | 0 | LA total emptying fraction | 0.583 | 0.106 | 0.286 |
| Current diabetes | Heart | LV chamber | 4 | 0 | 2 | LV mass index | −0.472 | 0.0372 | 0.0792 |
| Current diabetes | Heart | LV strain | 3 | 0 | 0 | LV circumferential strain | −0.261 | 0.337 | 0.541 |
| Current diabetes | Heart | RV strain | 3 | 0 | 0 | RV circumferential strain | −0.412 | 0.122 | 0.199 |
| HbA1c | Brain | Area | 220 | 0 | 2 | Right S calcarine surface area | 0.197 | 0.0176 | 1.00 |
| HbA1c | Brain | Mean curvature | 216 | 0 | 22 | Left insula mean curvature | 0.263 | 0.00407 | 0.250 |
| HbA1c | Brain | Cortical thickness | 216 | 0 | 16 | Right G temp sup G T transv thickness | −0.318 | 0.00493 | 0.441 |
| HbA1c | Brain | Thickness variability | 216 | 0 | 19 | Right G oc temp med Lingual thickness SD | −0.312 | 0.00317 | 0.469 |
| HbA1c | Brain | Volume | 216 | 0 | 30 | Left G front inf Orbital volume | −0.275 | 0.00122 | 0.263 |
| HbA1c | Heart | Atrial | 3 | 0 | 0 | LA total emptying fraction | 0.126 | 0.539 | 0.897 |
| HbA1c | Heart | LV chamber | 4 | 1 | 2 | LVEDVi | −0.219 | 0.00783 | 0.0313 |
| HbA1c | Heart | LV strain | 3 | 0 | 0 | LV radial strain | 0.157 | 0.114 | 0.232 |
| HbA1c | Heart | RV strain | 3 | 0 | 1 | RV radial strain | 0.157 | 0.0332 | 0.0997 |

Each row summarizes the modifier × APOE ε4 interaction tests within one phenotype family; FDR correction was applied separately within each modifier × domain × phenotype family. Current diabetes excludes Examination 8 code 2 ("yes, not now"). The only FDR-significant result was HbA1c × APOE ε4 for LVEDVi (followed up in Table S8). Destrieux parcels retain atlas abbreviations (G, gyrus; S, sulcus).

Abbreviations: APOE, apolipoprotein E; FDR, false discovery rate; HbA1c, glycated hemoglobin; LA, left atrial; LV, left ventricular; RV, right ventricular.

#### Table S10. Biological processes enriched in implicated cortical regions in the Allen Human Brain Atlas reference data

| Region | Heart–brain association | GO Biological Process | Enrichment ratio | P | FDR q | Overlap genes |
|---|---|---|---|---|---|---|
| Left middle temporal | LVEF × APOE ε4 → volume | Regulation Of Epithelial To Mesenchymal Transition (GO:0010717) | 4.06 | $6.50\times10^{-6}$ | 0.00571 | CLASP2, DAG1, EPHA4, GLIPR2, IL1B, MAD2L2, OLFM1, PTK2, SFRP1, SMAD4, SPRED1, TGFB3, USF3, VASN |
| Left middle temporal | LVEF × APOE ε4 → volume | Nervous System Development (GO:0007399) | 1.94 | $9.39\times10^{-5}$ | 0.0413 | ADORA1, APLP1, CDH1, CHRM2, CYP46A1, DOC2A, DPYSL5, DRD2, ENC1, GPM6A, IL1RAPL2, KCNQ2, KIRREL3, LIG4, LPAR1, MAL, MET, MST1R, MYLIP, NELL1, NPTX1, NRXN2, OLFM1, PAX8, PRKN, PRPS1, RNF103, SCN2A, SH3GL3, SOX10, SOX8, SPG7, TYRO3, VAX2, VLDLR, WDR62, ZC4H2 |
| Right superior frontal | LVESVi × APOE ε4 → surface area | Calcium-Ion Regulated Exocytosis (GO:0017156) | 6.66 | $1.36\times10^{-5}$ | 0.00765 | DOC2B, RAB3A, RIMS1, SYT1, SYT4, SYT5, SYT9, VAMP7 |
| Right superior frontal | LVESVi × APOE ε4 → surface area | Regulation Of Regulated Secretory Pathway (GO:1903305) | 5.62 | $1.86\times10^{-5}$ | 0.00765 | DOC2B, RAB15, RAB3A, RIMS1, RPH3AL, SYT1, SYT4, SYT5, SYT9 |
| Right superior frontal | LVESVi × APOE ε4 → surface area | Regulation Of Heterotypic Cell-Cell Adhesion (GO:0034114) | 8.00 | $5.21\times10^{-5}$ | 0.0143 | FLOT1, FLOT2, MAP2K5, MYADM, SKAP1, THY1 |
| Right superior frontal | LVESVi × APOE ε4 → surface area | Cholesterol Biosynthetic Process (GO:0006695) | 5.60 | $1.66\times10^{-4}$ | 0.0342 | CYP51A1, DHCR24, DHCR7, INSIG1, MSMO1, SC5D, TM7SF2 |
| Right superior frontal | LVESVi × APOE ε4 → surface area | Cellular Response To Calcium Ion (GO:0071277) | 3.49 | $2.59\times10^{-4}$ | 0.0428 | AKR1C3, CHP2, CPNE1, ECT2, NEUROD2, PPIF, SMPD1, SYT1, SYT4, SYT5, SYT9 |
| Right superior frontal | LVESVi × APOE ε4 → surface area | Diterpenoid Metabolic Process (GO:0016101) | 6.00 | $3.26\times10^{-4}$ | 0.0447 | AKR1C1, AKR1C3, AKR1C4, E2F1, LRP2, RDH12 |

Enrichment of genes characteristic of each implicated Desikan–Killiany region in the AHBA-derived regional reference atlas (adult postmortem microarray data; not Framingham participant expression). Only FDR-significant processes are listed; no process met q<0.05 for the right lingual region. Regional enrichment is distinct from the whole-cortex spatial-correspondence tests in Data S2 and does not establish mediation. See also Figure S5.

Abbreviations: AHBA, Allen Human Brain Atlas; APOE, apolipoprotein E; FDR, false discovery rate; GO, Gene Ontology; LVEF, LV ejection fraction; LVESVi, LV end-systolic volume index.

#### Table S11. Summary of molecular analyses for the four regional ventricular cardiac × APOE ε4 associations

| Quantity | LVEF × APOE ε4 → left middle temporal volume | LVESVi × APOE ε4 → right superior frontal surface area | LV mass index × APOE ε4 → right lingual volume | LV mass index × APOE ε4 → right lingual surface area |
|---|---|---|---|---|
| Primary interaction β | 0.233 | −0.170 | 0.305 | 0.226 |
| Primary 95% CI | 0.087 to 0.380 | −0.260 to −0.081 | 0.144 to 0.467 | 0.095 to 0.357 |
| Primary P | 0.00176 | $1.97\times10^{-4}$ | $2.16\times10^{-4}$ | $7.09\times10^{-4}$ |
| Primary FDR q | 0.0352 | 0.00433 | 0.00604 | 0.0198 |
| WGS pathways tested | 7 | 7 | 7 | 7 |
| Top-ranked WGS pathway | mitochondrial energetics | insulin glucose | insulin glucose | cardiac remodeling fibrosis |
| Top-ranked WGS omnibus P | 0.365 | 0.319 | 0.0841 | 0.396 |
| Top-ranked WGS FDR q | 0.896 | 0.896 | 0.896 | 0.896 |
| Targeted AHBA pathways tested | 7 | 7 | 7 | 7 |
| Top-ranked targeted AHBA pathway | mitochondrial energetics | mitochondrial energetics | endothelial vascular | endothelial vascular |
| Top-ranked targeted AHBA Spearman ρ | 0.171 | −0.141 | 0.180 | 0.221 |
| Top-ranked targeted AHBA spatial P | 0.452 | 0.530 | 0.333 | 0.319 |
| Targeted AHBA FDR q | 0.914 | 0.914 | 0.914 | 0.914 |
| GO/Reactome discovery tests | 4,417 | 4,417 | 4,417 | 4,417 |
| Top-ranked discovery library | Reactome | Reactome | Reactome | Reactome |
| Top-ranked discovery term | APC Truncation Mutants Have Impaired AXIN Binding R-HSA-5467337 | RAF Activation R-HSA-5673000 | SUMOylation Of Transcription Factors R-HSA-3232118 | Frs2-mediated Activation R-HSA-170968 |
| Top-ranked discovery Spearman ρ | 0.564 | −0.576 | 0.502 | −0.400 |
| Top-ranked discovery spatial P | $4.00\times10^{-4}$ | $6.00\times10^{-4}$ | 0.00100 | $2.00\times10^{-4}$ |
| Top-ranked discovery within-map/library q | 0.155 | 0.432 | 0.955 | 0.130 |
| Top-ranked discovery global q | 0.604 | 0.604 | 0.604 | 0.604 |
| Global FDR-significant discovery terms | 0 | 0 | 0 | 0 |
| Mechanistic triangulation conclusion | No convergence | No convergence | No convergence | No convergence |

WGS: participant-level TOPMed whole-genome sequencing; seven gene-set groups tested for modification of each association (28 omnibus tests, FDR across all 28). Targeted AHBA: Spearman correlation between seven pathway-expression maps and each interaction map across 56 Desikan–Killiany parcels, with P values from 5,000 Moran spatial surrogates and FDR across 28 comparisons (Data S2). Discovery: GO Biological Process and Reactome terms, with q values within each map and library and across all 17,668 tests (global).

"No convergence": no corrected association across WGS, targeted AHBA spatial testing, or GO/Reactome spatial discovery.

Abbreviations: AHBA, Allen Human Brain Atlas; APOE, apolipoprotein E; CI, confidence interval; FDR, false discovery rate; GO, Gene Ontology; WGS, whole-genome sequencing.

### Supplementary Figures

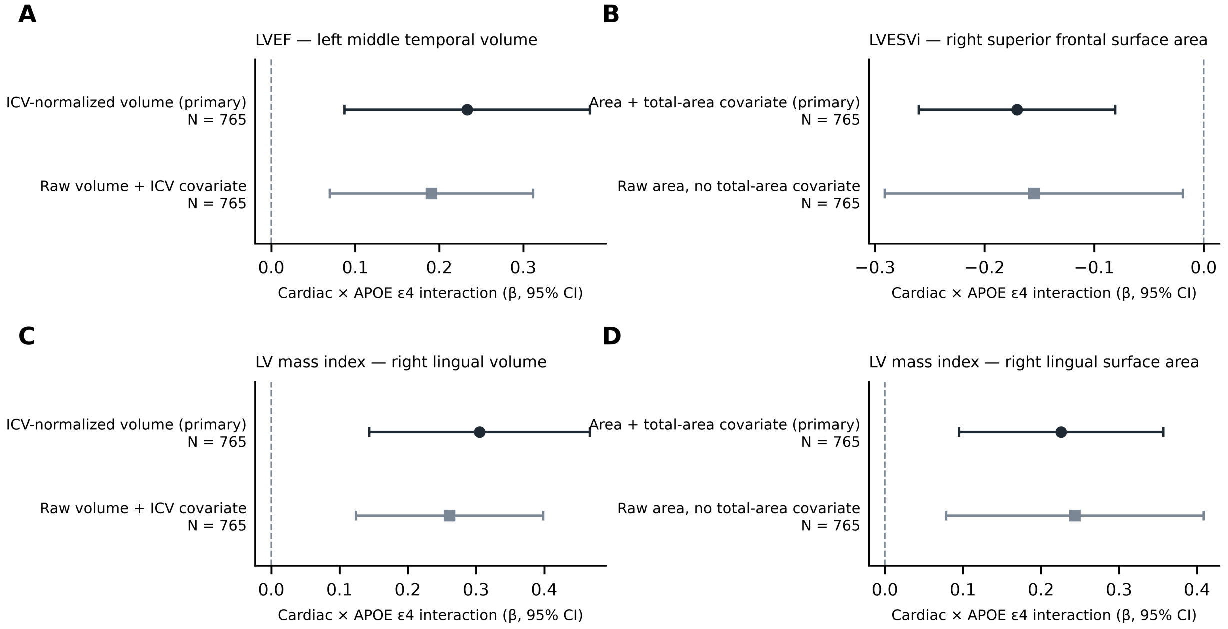


**Figure S1. Alternative morphometric specifications of the four regional ventricular interactions.** Cardiac × APOE ε4 interaction coefficients (95% CI) for the primary specification and an alternative specification: raw regional volume with intracranial volume as a covariate (volumes) or raw surface area without the total-cortical-area covariate (surface areas); N=765. Values are in Supplementary Data S2.

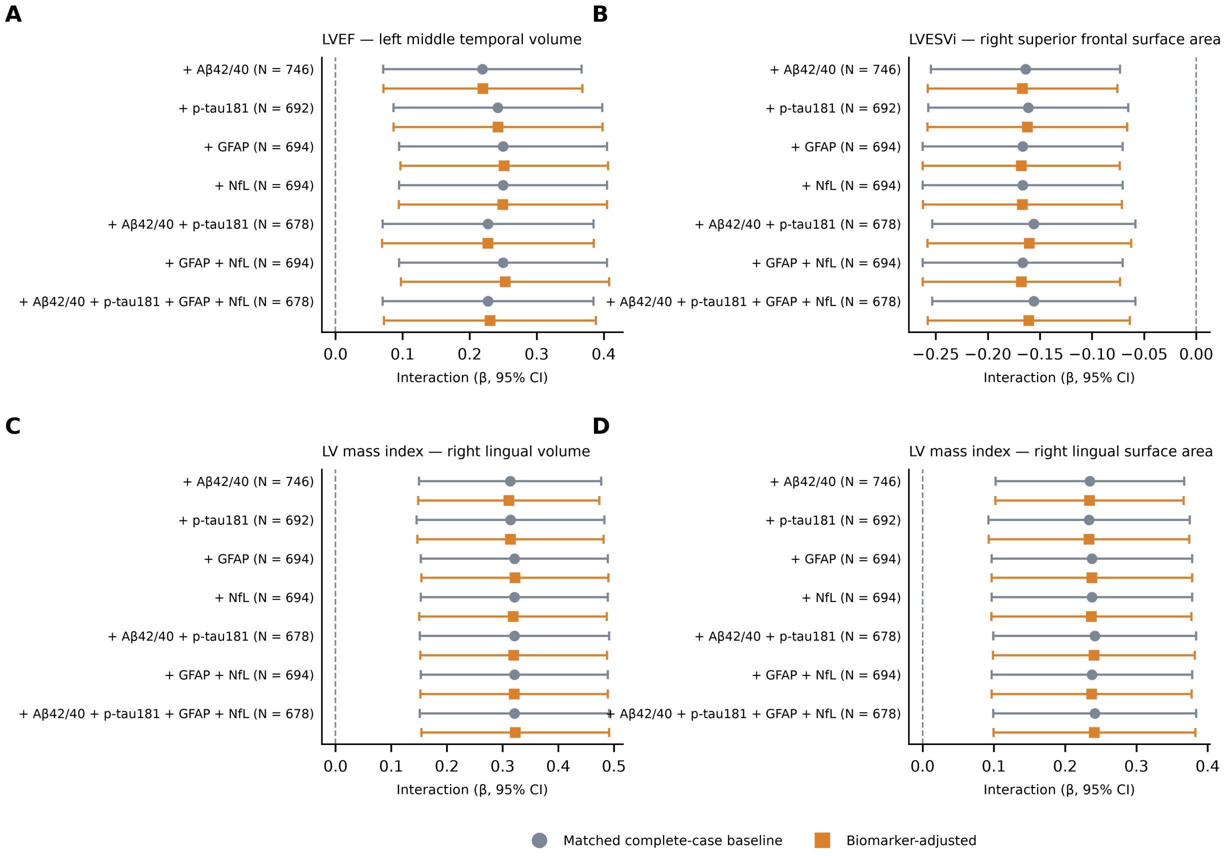


**Figure S2. Plasma-biomarker adjustment of the four regional ventricular interactions.** Interaction coefficients (95% CI) in matched complete-case baseline models (grey) and after adjustment (orange) for Aβ42/40, p-tau181, GFAP, NfL, Aβ42/40 + p-tau181, GFAP + NfL, or all four biomarkers (N=678–746). Absolute changes in interaction magnitude were at most 3.2%. Values are in Supplementary Data S2.

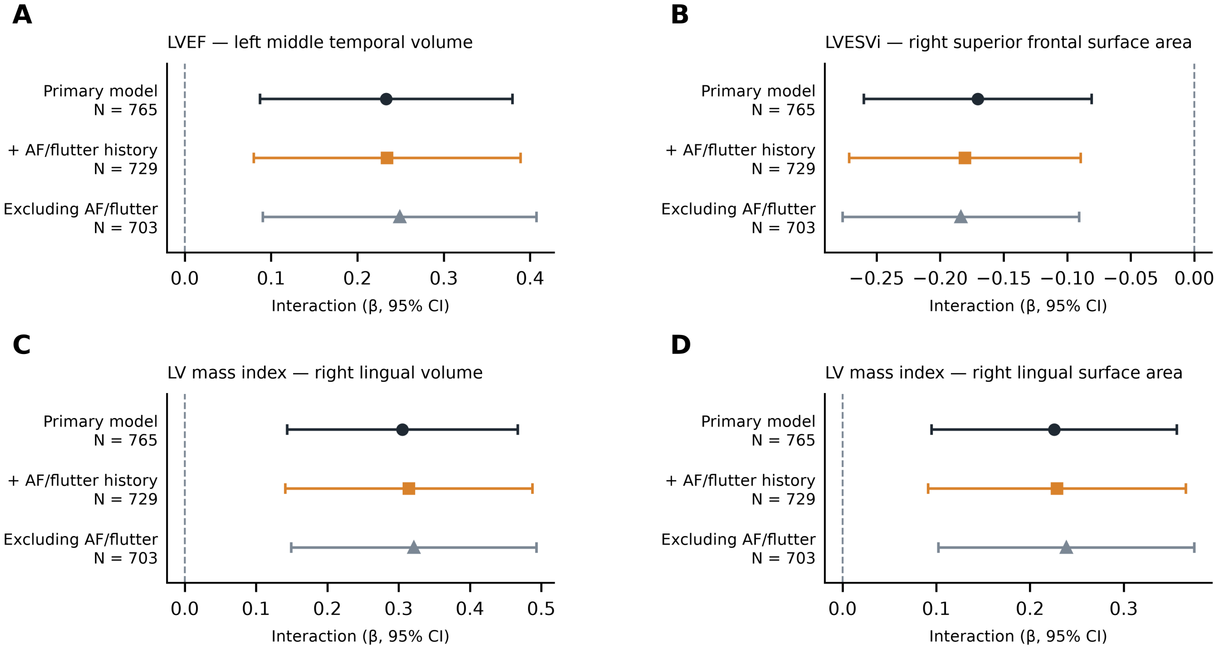


**Figure S3. Atrial fibrillation/flutter sensitivity of the four regional ventricular interactions.** Interaction coefficients (95% CI) in the primary model, after adjustment for AF/flutter history, and after excluding participants with AF/flutter. Values are in Supplementary Data S2.

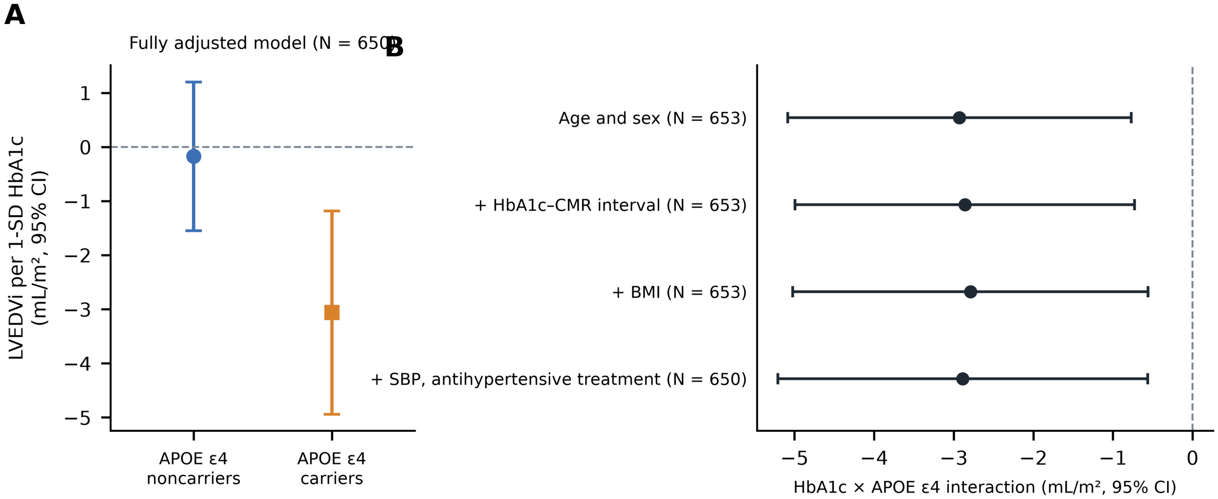


**Figure S4. HbA1c × APOE ε4 and LV end-diastolic volume index (exploratory).** (A) Change in LVEDVi per 1-SD higher HbA1c (95% CI) among APOE ε4 noncarriers and carriers in the fully adjusted model. (B) The HbA1c × APOE ε4 interaction across nested models (Table S8).

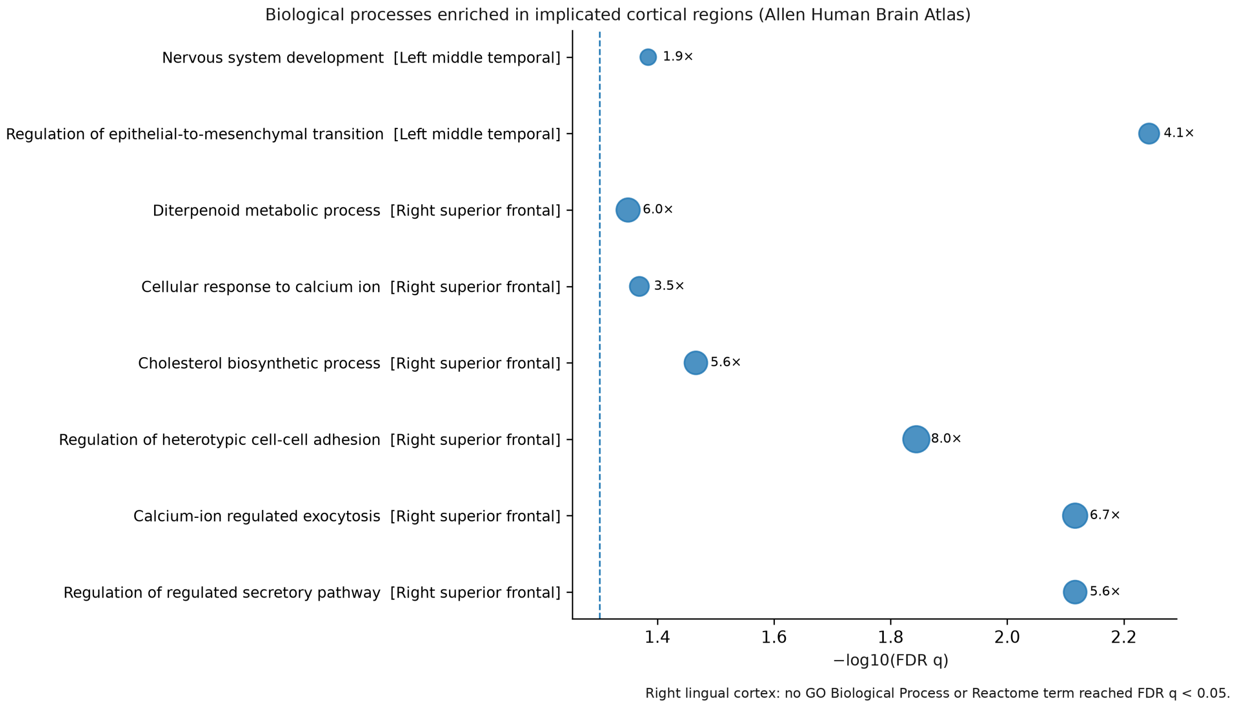


**Figure S5. Biological processes enriched in implicated cortical regions in Allen Human Brain Atlas reference data.** Enrichment of Gene Ontology biological processes among genes characteristic of the left middle temporal, right superior frontal, and right lingual regions, relative to other cortical regions (Table S10). This descriptive reference-atlas analysis characterizes regions selected from the exploratory regional screen; whole-cortex spatial-correspondence tests are summarized in Table S11 and reported in full in Supplementary Data S2.

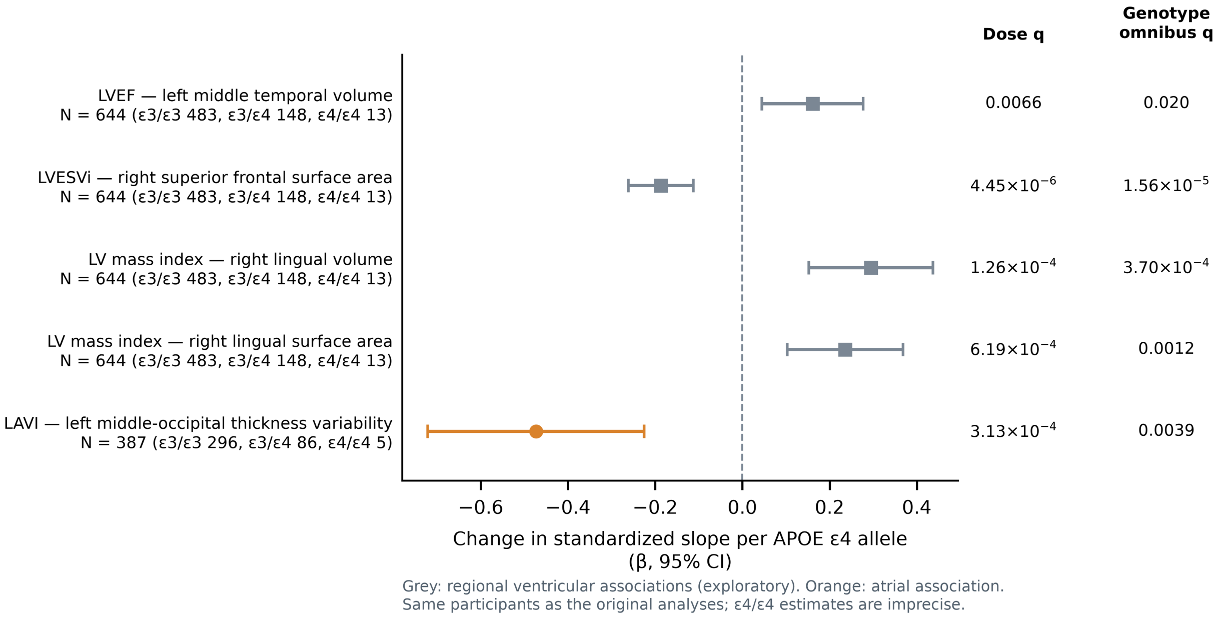


**Figure S6. APOE ε4 allele-dose sensitivity analyses of the regional cardiac–brain associations.** Change in the standardized cardiac–brain slope per additional ε4 allele (95% CI) in ε3/ε3, ε3/ε4, and ε4/ε4 participants, with dose and categorical-genotype q values (Table 3). Grey, regional ventricular associations (exploratory); orange, atrial association. These analyses use the participants in whom the associations were identified and are not independent confirmation; ε4/ε4 estimates are imprecise.
